# Quantitative plasma proteomics of survivor and non-survivor COVID-19 patients admitted to hospital unravels potential prognostic biomarkers and therapeutic targets

**DOI:** 10.1101/2020.12.26.20248855

**Authors:** Daniele C. Flora, Aline D. Valle, Heloisa A. B. S. Pereira, Thais F. Garbieri, Nathalia R. Buzalaf, Fernanda N. Reis, Larissa T. Grizzo, Thiago J. Dionisio, Aline L. Leite, Virginia B. R. Pereira, Deborah M. C. Rosa, Carlos F. Santos, Marília A. Rabelo Buzalaf

## Abstract

The development of new approaches that allow early assessment of which cases of COVID-19 will likely become critical and the discovery of new therapeutic targets are urgent demands. In this cohort study, we performed proteomic and laboratorial profiling of plasma from 163 patients admitted to Bauru State Hospital (Bauru, SP, Brazil) between May 4^th^ and July 4^th^, 2020, who were diagnosed with COVID-19 by RT-PCR nasopharyngeal swab samples. Plasma samples were collected upon admission for routine laboratory analyses and shotgun quantitative label-free proteomics. Based on the course of the disease, the patients were further divided into 3 groups: a) mild symptoms, discharged without admission to an intensive care unit (ICU) (n=76); b) severe symptoms, discharged after admission to an ICU (n=56); c) critical, died after admission to an ICU (n=31). White cells and neutrophils were significantly higher in severe and critical patients compared to mild ones. Lymphocytes were significantly lower in critical patients compared to mild ones and platelets were significantly lower in critical patients compared to mild and severe ones. Ferritin, TGO, urea and creatinine were significantly higher in critical patients compared to mild and severe ones. Albumin, CPK, LDH and D-dimer were significantly higher in severe and critical patients compared to mild ones. PCR was significantly higher in severe patients compared to mild ones. Proteomic analysis revealed marked changes between the groups in plasma proteins related to complement activation, blood coagulation, antimicrobial humoral response, acute inflammatory response, and endopeptidase inhibitor activity. Higher levels of IREB2, GELS, POLR3D, PON1 and ULBP6 upon admission to hospital were found in patients with mild symptoms, while higher levels of Gal-10 were found in critical and severe patients. This needs to be validated in further studies. If confirmed, pathways involving these proteins might be potential new therapeutic targets for COVID-19.

## Introduction

In December 2019, in Wuhan, China, several patients were diagnosed with pneumonia caused by a new beta-coronavirus, initially called 2019-nCoV and later given the official name of severe acute respiratory syndrome coronavirus 2 (SARS-CoV-2) (1). The virus rapidly spread all over the world originating an unprecedented pandemic. Within one year, about 80 million individuals were infected, leading to nearly 1.8 million deaths globally.

Nearly 80% of patients affected by COVID-19 have only mild symptoms, recovering with conventional medical treatment, or even without any treatment (2, 3). Around 20% of affected patients, however, develop respiratory distress, requiring oxygen therapy or even mechanical ventilation and nearly 10% of them need admission to intensive care units (ICUs) (4). Moreover, mortality of late-stage acute respiratory distress syndrome (ARDS) in consequence of COVID-19 is remarkably high. Around 48-90% of patients intubated and placed on mechanical ventilation do not survive, a percentage significantly higher than that associated with intubation for other viral pneumonias, which is nearly 22% (5, 6). In addition, for non-survivors, median duration from admission to hospital to death is 10 days (6). Considering the high virulence of SARS-CoV-2, the lack of effective therapies, the high rates of mortality among critical patients, as well as the short period between admission to hospital and death, the development of new approaches that allow early assessment of which cases will likely become critical, as well as the discovery of new therapeutic targets are crucial.

Alterations of plasma proteins are good indicators of pathophysiological changes caused by several diseases, including viral infections. In this sense, plasma proteomics is widely used for biomarker discovery (7). Only a few studies so far have performed proteomic profiling of plasma/serum of COVID-19 patients (8-12). Among them, some enrolled only a few COVID-19 patients (9, 11, 12), while another one enrolled patients with no need of hospitalization that were compared with hospitalized ones, without distinguishing the severity of the disease among hospitalized patients (8). The study by Overmyer et al. (10) evaluated plasma samples of 102 and 26 patients who tested positive and negative for SARS-CoV-2, respectively. These samples were collected from April 6^th^, 2020 through May 1^st^, 2020 from patients admitted to Albany Medical Center in Albany, NY, USA, presenting severe respiratory symptoms probably related to COVID-19. The positive patients were divided into two severity groups, based on admission to an ICU or not. The authors identified proteins and metabolites offering pathophysiological insights of the disease, as well as offered therapeutic suggestions. The main limitation, however, was the lack of association of omics data with survival, which is the most remarkable outcome measure (10). Here we profiled the host responses to SARS-CoV-2 by performing shotgun label-free quantitative proteomic analysis of plasma samples from a cohort of 163 COVID-19 patients admitted to Bauru State Hospital (HEB), Brazil, between May 4^th^, 2020 and July 4^th^, 2020. Plasma samples were collected upon admission and patients were divided into 3 groups, based on the course of the disease, including both survivors (mild and severe patients) and non-survivors (critical patients). The proteomic findings were associated with the severity of the disease, as well as with the laboratory findings.

## Methods & Materials

### 2.1 Ethical aspects

This project was approved by the Ethical Committee of Bauru School of Dentistry, University of São Paulo (CAAE 31019820.8.0000.5417) upon acquiescence of the Nucleus of Teaching and Research of the HEB. Patients participated after they (or their relatives) signed an informed consent document.

### 2.2 Study design and patients

**Figure 1** illustrates the main characteristics of the study that had a cohort design. All patients admitted to the HEB (Bauru, SP, Brazil) between May 4^th^ and July 4^th^, 2020, who were diagnosed with COVID-19 by RT-PCR nasopharyngeal swab samples and provided a signed informed consent document were included. Exclusion criterium was refusal to participate in the study by the patient or relative.

**Figure 1.**
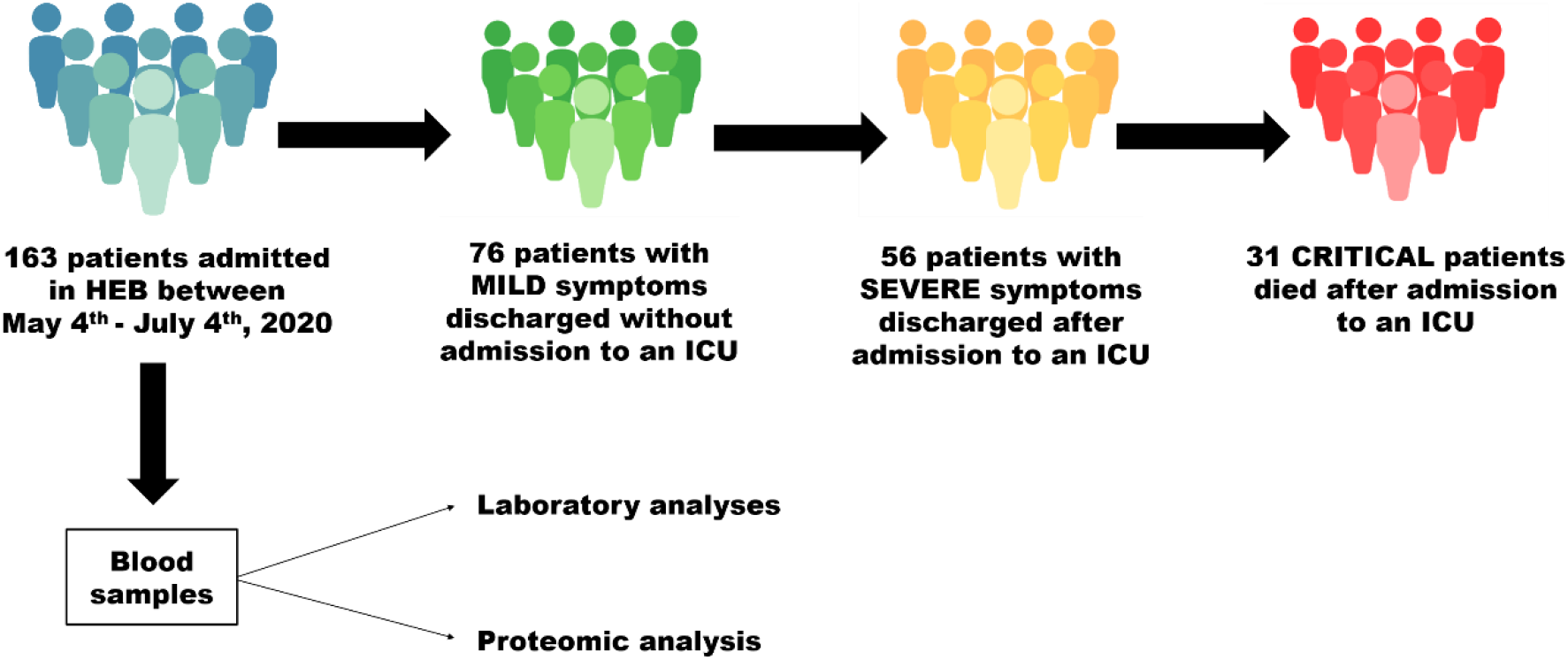
Experimental design of the study.

All patients received the basic routine support established by the hospital that included oxygen support, invasive and non-invasive mechanic ventilation, use of antibiotics, use of vasopressor, use of anticoagulant, renal support therapy and use of corticoid if necessary.

### 2.3. Comparisons and sampling

The patients were divided into 3 groups, based on the course of the disease, as follows: a) patients with mild symptoms that were discharged without admission to an ICU; b) patients with severe symptoms that were discharged after admission to an ICU; c) critical patients, who were admitted to an ICU and died (**Figure 1**).

At admission, blood samples were collected for routine laboratory analyses at the hospital, as follows: white blood cell count, neutrophil count, lymphocyte count, eosinophil count, platelet count, hemoglobin, red blood cell count, ferritin, albumin, aspartate aminotransferase (TGO), alanine aminotransferase (TGP), creatinine phosphokinase (CPK), urea, creatinine, C-reactive protein (PCR), lactate dehydrogenase (LDH) and D-dimer. Shortly after collection, an aliquot of blood samples was centrifuged at 2000 x g for 10 min and the plasma fraction was stored at −80°C for proteomic analysis.

### 2.4 Preparation of the plasma samples for proteomic analysis

Initially, the samples were submitted to depletion, as previously described (13). For this, 60 μL of plasma were diluted in 180 μL buffer A (Equil / Load / Wash; Agilent) and vortexed. In sequence, the solution was loaded on Filter Spin (0.22 um; Agilent) and centrifuged at 16,000 g for 1 min, collected in a tube and then followed to Multiple Affinity Removal Column (Agilent Technologies), according to the manufacturer’s protocol. Buffer A (Agilent) was used for washing and balancing column, while Buffer B (Agilent) was employed for elution of the bound proteins from the column. The low-abundance flow-through fraction was collected and stored at –20 ° C until analyses.

In order that the samples followed to proteomic analyses, it was necessary to exchange the depletome buffer with 50 nM ammonium bicarbonate, using Amicon® Ultra 4 mL Centrifugal Filters (Merck). The final volume was 600 µL. The same volume of urea solution (8 mM Urea in 50 nM ammonium bicarbonate buffer) was added. Samples were then quantified (14) and a volume corresponding to 100 μg proteins was reduced with dithiothreitol (100 mM, 40°C, 30 min) and alkylated with iodoacetamide (300 mM, 30 min, room temperature, in the dark). Then, samples were digested with Pierce™ Trypsin

Protease, MS Grade (Thermo Scientific) at a ratio of 2: 100 (w / w trypsin / protein) for 14 h to 16 h at 37°C. Digestion was quenched with 5% trifluoroacetic acid (TFA) for 15 min at room temperature. The samples were centrifuged at 14,000 g, 6°C for 30 min. The supernatant was recovered, and the samples were purified and concentrated using Pierce C18 Spin Columns (Thermo Scientific). In sequence, samples were quantified (14), dried and stored for proteomic analyses..

### 2.5 Shotgun label free quantitative proteomic analysis

The analysis was performed in a nanoACQUITY UPLC system (Waters, Milford, MA) coupled to a Xevo Q-TOF G2 mass spectrometer (Waters, Milford, MA), as previously described (15).

The raw data were processed with ProteinLynx Global Server (PLGS) 3.0.3. Data were extracted, aligned, and searched against the Uniprot human proteomic database, version 2020-01, appended with the enolase (*S. Cerevisae*) sequence. The ion accounting algorithm used has been described previously (16). PLGS utilizes the drift time of mobility separated peptides to increase the specificity of alignment/association for precursor and product ions. PLGS also assigns peptide identifications to proteins through an iterative matching process (16). Data were further processed with Microsoft Excel for additional data analysis and for the generation of figures and tables.

Label-free proteomic quantification was performed using PLGS software. Difference in expression among the groups was calculated using Monte-Carlo algorithm embedded in the software and expressed as p<0.05 for proteins present in lower abundance and 1-p>0.95 for proteins present in higher abundance.

The software CYTOSCAPE 3.7.2 (JAVA) was used to build networks of molecular interaction between the identified proteins, with the aid of ClueGo and ClusterMarker applications.

### 2.6 Statistical analysis

The software GraphPad InStat (version 3.0 for Windows; GraphPad Software Inc. La Jolla, Ca, USA) was used. Data were checked for normal distribution using Kolmogorov-Smirnov test and for homogeneity using Bartlett’s test for the selection of the appropriate statistical test. The significance level, in all cases, was set at 5%.

## Results

### 3.1 Characterization of the patients included in the study

The number of patients included in each group was 76, 56 and 31 for mild, severe and critical patients, respectively, totaling 163 patients (82 men and 81 women). The median age of critical patients (73.0 years) was significantly higher than that in mild (51.0 years) and severe (56.5 years) patients that did not significantly differ from each other (Table 1). Critical patients were significantly older than mild and severe ones, whose ages did not significantly differ. The percentages of females/males were 53.9/46.1, 53.6/46.4, and 35.5/64.5 for mild, severe and critical patients, respectively. The characteristics of the patients in each of the groups regarding comorbidity and lifestyle conditions are summarized in **Table 1**.

**Table 1.** Characteristics* of the patients admitted to Bauru State Hospital, Brazil, between May 4^th^ and July 4^th^, 2020, who were diagnosed with COVID-19 by RT-PCR nasopharyngeal swab samples

| Characteristics | Mild | Severe | Critical | p |
| --- | --- | --- | --- | --- |
| *Median age, years (95% CI)* | 51.0 (48.8-56.7)<sup>a</sup> | 56.5 (51.8-60.4)<sup>a</sup> | 73.0 (63.7-72.7)<sup>b</sup> | <0.01 |
| *Female (n, %)* | 41, 53.9% | 30, 53.6% | 11, 35.5% |  |
| *Male (n, %)* | 35, 46.1% | 26, 46.4% | 20, 64.5% |  |
| *Comorbidity* |  |  |  |  |
| Hypertension | 31.6% | 49.1% | 64.5% |  |
| Diabetes | 21.1% | 41.8% | 32.3% |  |
| Cardiovascular disease | 10.5% | 10.9% | 9.7% |  |
| Obesity | 22.4% | 14.5% | 3.2% |  |
| COPD | 5.3% | 10.9% | 12.9% |  |
| Cancer | 2.6% | 5.5% | 3.2% |  |
| Nephropathy | 3.9% | 5.5% | 16.1% |  |
| Hepatic disease | 1.3% | 0 | 0 |  |
| Stroke | 5.3% | 1.8% | 6.5% |  |
| Autoimmune disease | 0 | 0 | 6.5% |  |
| *Lifestyle* |  |  |  |  |
| Smoking | 3.9% | 12.7% | 3.2% |  |
| Drinking | 2.6% | 3.6% | 0 |  |

\* at admission.
Kruskal-Wallis and Dunn's test. n=76 (Mild), 56 (Severe) and 31 (Critical).
COPD – Chronic obstructive pulmonary disease.
Patients with mild symptoms were discharged without admission to an intensive care unit (ICU)
Patients with severe symptoms were discharged after admission to an ICU
Critical patients died after admission to an ICU.

### 3.2. Laboratory findings

For full blood counts, no significant differences were found among the groups for red cells, hemoglobin and eosinophils. White cells and neutrophils were significantly higher in severe and critical patients compared to mild ones. On the other hand, lymphocytes were significantly lower in critical patients compared to mild ones and platelets were significantly lower in critical patients compared to mild and severe ones. The other differences were not significant (p>0.05) (**Table 2**).

**Table 2.** Laboratory variables^&^ of patients admitted to Bauru State Hospital, Brazil, between May 4^th^ and July 4^th^, 2020, who were diagnosed with COVID-19 by RT-PCR nasopharyngeal swab samples

| Characteristics | Mild | Severe | Critical | p |
| --- | --- | --- | --- | --- |
| *Full blood counts* |  |  |  |  |
| Red blood cell, X10<sup>6</sup>/mm<sup>3</sup> | 4.43±0.58<sup>a</sup> | 4.24±0.69<sup>a</sup> | 4.12±0.79<sup>a</sup> | 0.058** |
| White blood cell, /mm<sup>3</sup> | 5930 (5565-6949)<sup>a</sup> | 8050 (7090-9090)<sup>b</sup> | 8020 (7349-12024)<sup>b</sup> | <0.05* |
| Neutrophil, /mm<sup>3</sup> | 4112 (4086-5707)<sup>a</sup> | 6018 (5553-7628)<sup>b</sup> | 5849 (5613 -9889)<sup>b</sup> | <0.05* |
| Lymphocyte, /mm<sup>3</sup> | 954 (986-1397)<sup>a</sup> | 870 (817 – 1353)<sup>ab</sup> | 529 (458-1255)<sup>b</sup> | <0.01* |
| Hemoglobin, g/dL | 13.0 (12.5-13.8)<sup>a</sup> | 12.8 (12.2-13.2)<sup>a</sup> | 12.5 (11.5-13.2)<sup>a</sup> | 0.217* |
| Eosinophil, /mm<sup>3</sup> | 0 (15.4-51.8)<sup>a</sup> | 0 (11.7-104.6)<sup>a</sup> | 0 (7.7-61.2)<sup>a</sup> | 0.848* |
| Platelet, X10<sup>3</sup>/mm<sup>3</sup> | 220 (210-245)<sup>a</sup> | 220 (207-268)<sup>a</sup> | 176 (154-245)<sup>b</sup> | <0.05* |
| *Biochemical tests* |  |  |  |  |
| Ferritin, µg/L | 417 (511-796)<sup>a</sup> | 631 (664-1100)<sup>a</sup> | 931 (883-1474)<sup>b</sup> | <0.01* |
| Albumin, g/dL | 3.60 (3.42-3.82)<sup>a</sup> | 3.20 (3.17-3.41)<sup>b</sup> | 3.1 (2.82-3.23)<sup>b</sup> | <0.01* |
| TGO, U/L | 29.5 (31.5-50.0)<sup>a</sup> | 37.0 (36.7-57.1)<sup>a</sup> | 46.0(40.9-81.5)<sup>b</sup> | <0.05* |
| TGP, U/L | 29.0 (33.5-51.1)<sup>a</sup> | 36.0 (37.6-65.6)<sup>a</sup> | 31.0 (27.8-61.8)<sup>a</sup> | 0.79* |
| CPK, U/L | 75 (85-138)<sup>a</sup> | 120 (173-344)<sup>b</sup> | 124 (140-865)<sup>b</sup> | <0.01* |
| Urea, mg/dL | 32.8 (34.3-50.6)<sup>a</sup> | 33.8 (35.1-54.5)<sup>a</sup> | 44.0 (43.3-60.9)<sup>b</sup> | <0.05* |
| Creatinine, mg/dL | 0.80 (1.00-1.99)<sup>a</sup> | 0.80 (0.86-1.35)<sup>a</sup> | 1.20 (1.19-4.60)<sup>b</sup> | <0.05* |
| PCR, mg/L | 46.0 (60.3-93.7)<sup>a</sup> | 120.6 (92.5-140.0)<sup>b</sup> | 102.0 (86.5-151.9)<sup>ab</sup> | <0.01* |
| LDH, U/L | 213 (221-263)<sup>a</sup> | 314 (285-363)<sup>b</sup> | 402 (290-531)<sup>b</sup> | <0.001* |
| D-dimer, mg/L | 0.73 (0.83-1.32)<sup>a</sup> | 1.11 (1.55-2.62)<sup>b</sup> | 2.14 (1.66-6.43)<sup>b</sup> | <0.01* |

<sup>&</sup> at admission.
\* Kruskal- Wallis and Dunn's tests. Data are expressed as median (95% CI).
\*\* ANOVA and Tukey's tests. Data are expressed as mean±SD.
Patients with mild symptoms were discharged without admission to an intensive care unit (ICU).
Patients with severe symptoms were discharged after admission to an ICU.
Critical patients died after admission to an ICU.

As for the biochemical tests, no significant differences among the groups were found for TGP levels. Ferritin, TGO, urea and creatinine were significantly higher in critical patients compared to mild and severe ones. Albumin, CPK, LDH and D-dimer were significantly higher in severe and critical patients compared to mild ones. PCR was significantly higher in severe patients compared to mild ones. The other differences were not significant (p>0.05) (**Table 2**).

### 3.3 Proteomic analysis

Figures 2, 3 and 4 show the functional classification according to the biological processes with the most significant term, for the comparisons severe *vs*. mild, critical *vs*. mild and critical *vs*. severe, respectively. For the comparison severe *vs*. mild, the categories with the highest percentages of associated genes were: Phagocytosis (25%), Blood coagulation (14%), Endopeptidase inhibitor activity (9%), Antimicrobial humoral response (7%) and Acute inflammatory response (7%) (**Figure 2**). Regarding the comparison between critical *vs*. mild, the most affected categories were: Complement activation (29%), Blood coagulation (13%), Endopeptidase inhibitor activity (9%), Antibacterial humoral response (7%) and Acute inflammatory response (7%) (**Figure 3**). As for the comparison critical *vs*. severe, the most affected categories were: Regulation of humoral immune response (22%), Blood coagulation (13%), Negative regulation of endopeptidase activity (12%), Antimicrobial humoral response (7%) and Acute inflammatory response (6%) (**Figure 4**).

**Figure 2.**
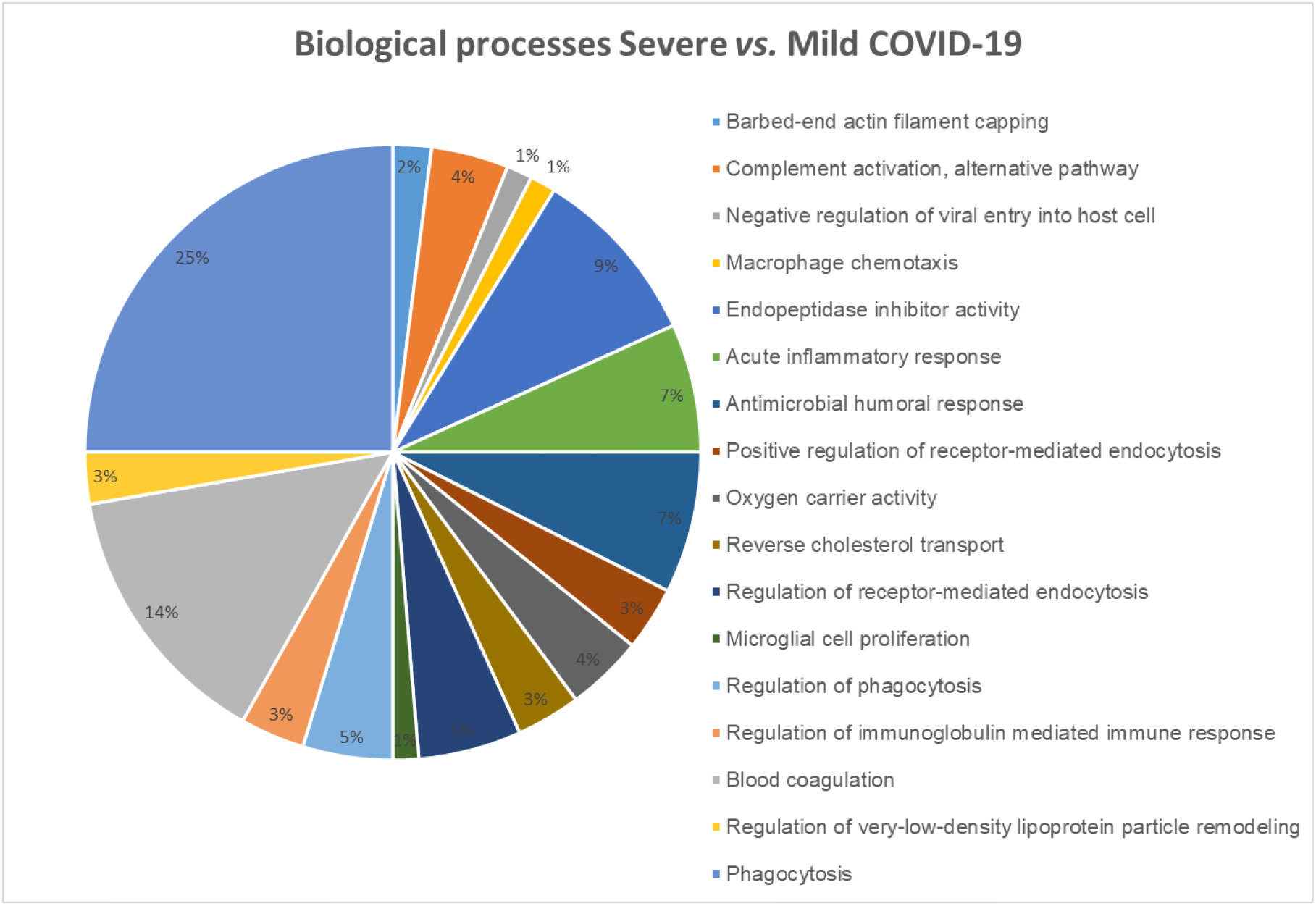
Functional distribution of proteins identified with differential expression in the plasma of patients admitted to Bauru State Hospital, Brazil, between May 4^th^ and July 4^th^, 2020, who were diagnosed with severe or mild COVID-19. Categories of proteins based on GO annotation Biological Process. Terms significant (Kappa=0.03) and distribution according to percentage of number of genes association

**Figure 3.**
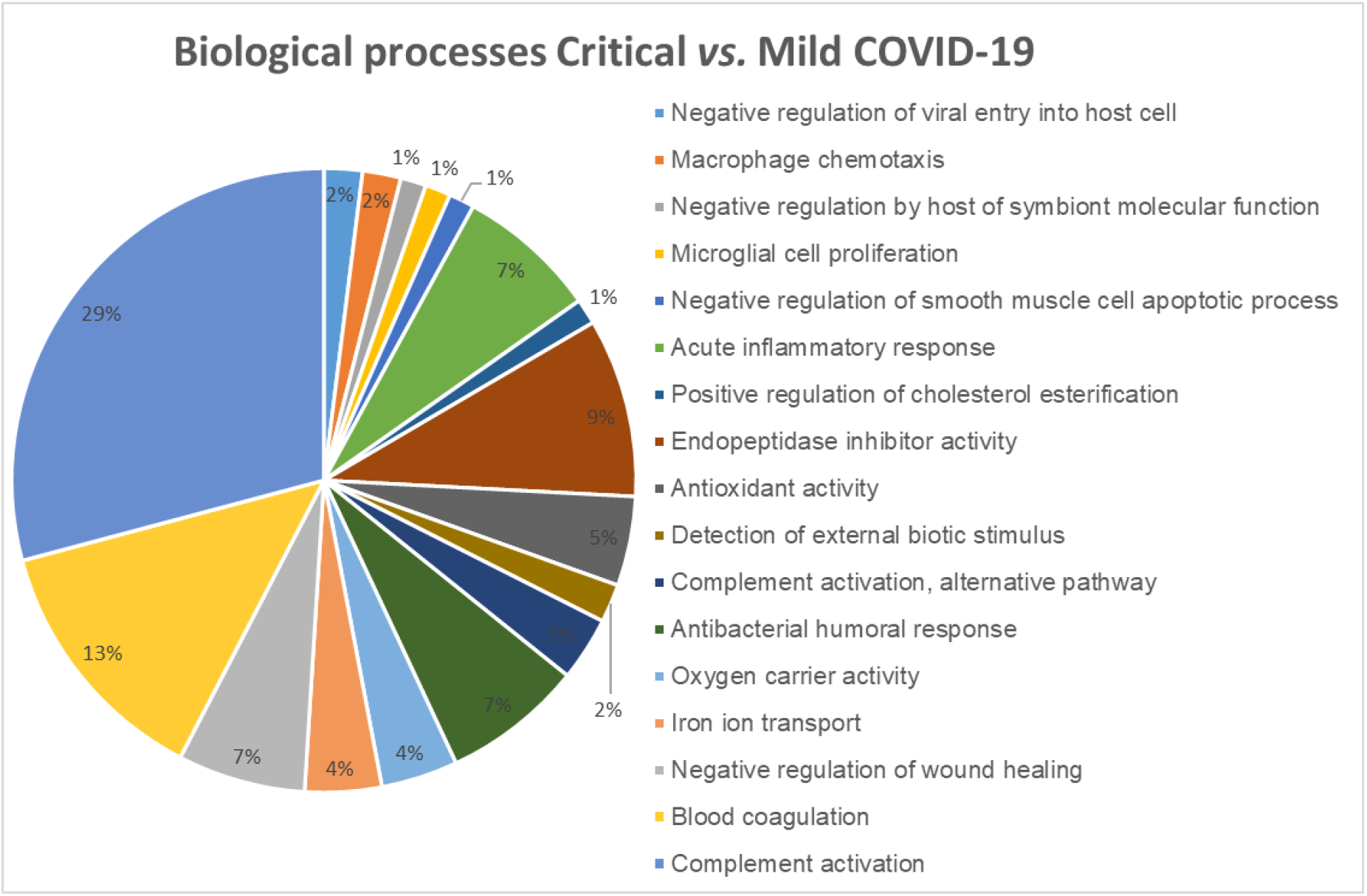
Functional distribution of proteins identified with differential expression in the plasma of patients admitted to Bauru State Hospital, Brazil, between May 4^th^ and July 4^th^, 2020, who were diagnosed with critical or mild COVID-19. Categories of proteins based on GO annotation Biological Process. Terms significant (Kappa=0.03) and distribution according to percentage of number of genes association

**Figure 4.**
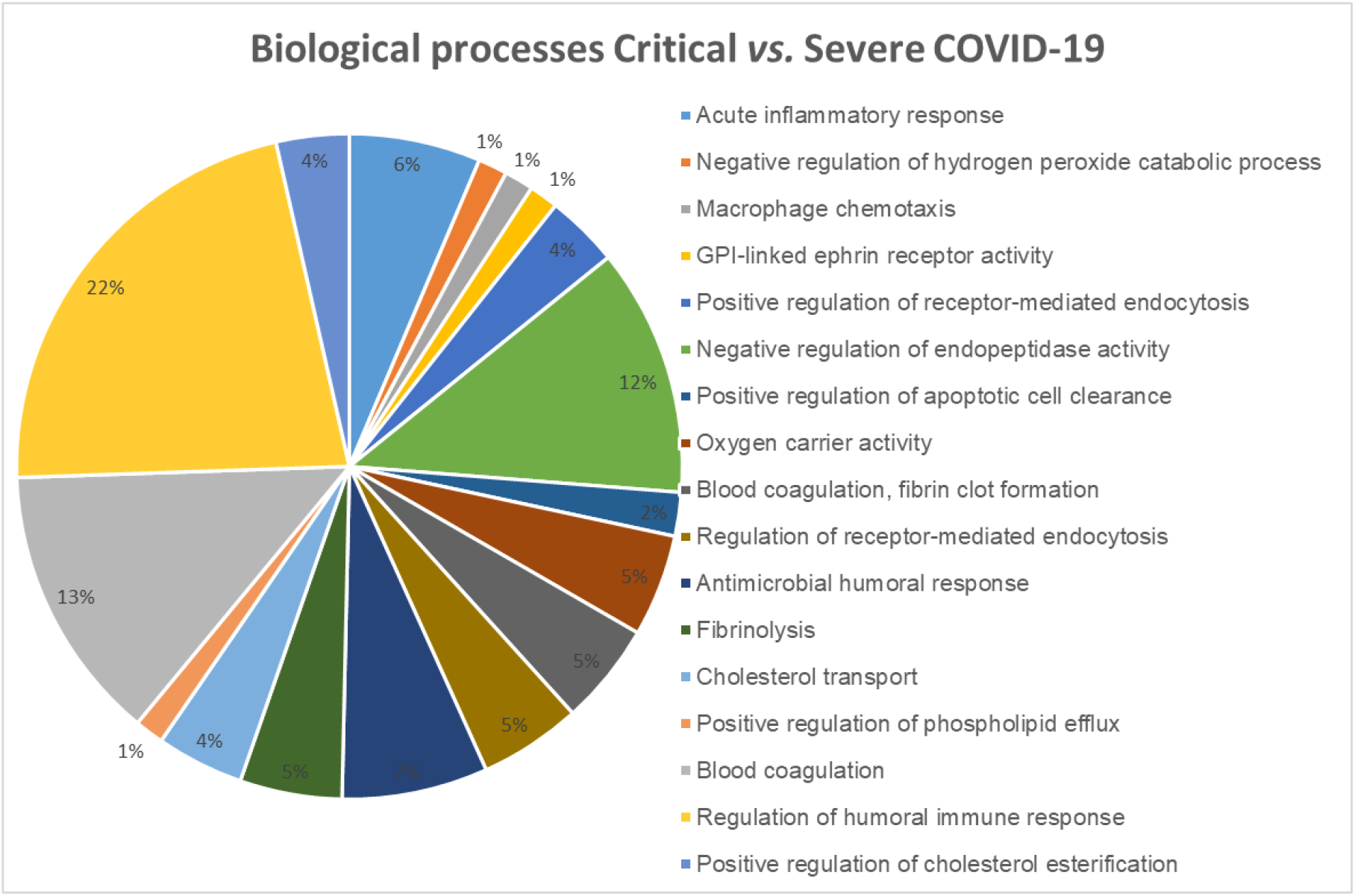
Functional distribution of proteins identified with differential expression in the plasma of patients admitted to Bauru State Hospital, Brazil, between May 4^th^ and July 4^th^, 2020, who were diagnosed with critical or severe COVID-19. Categories of proteins based on GO annotation Biological Process. Terms significant (Kappa=0.03) and distribution according to percentage of number of genes association

In the comparison between severe *vs*. mild patients (**Table 3**), 18 proteins were increased and 26 were exclusively found in the first. Proteins with the highest increases were Plasminogen-like protein (Q02325; 6.2-fold), Serum paraoxonase/arylesterase 1 (PON-1; P27169; 5.1-fold), Serum amyloid A-2 (P0DJI9; 3.2-fold) and A-1 (P0DJI8; 2.2-fold). Other increased proteins were C-reactive protein (CRP; P02741), Apoliprotein-C-III (P0265) and Alpha-1-antichymotrypsin (P01011). Among the proteins exclusively found in the patients with severe symptoms are those related to antimicrobial humoral response, such as Galectin-10 (Gal-10; Q05315), also known as Charcot-Leyden crystal (CLC) protein, and blood coagulation, such as C4b-binding protein beta chain (P20851) and Coagulation factor XII (P00748).

**Table 3.**
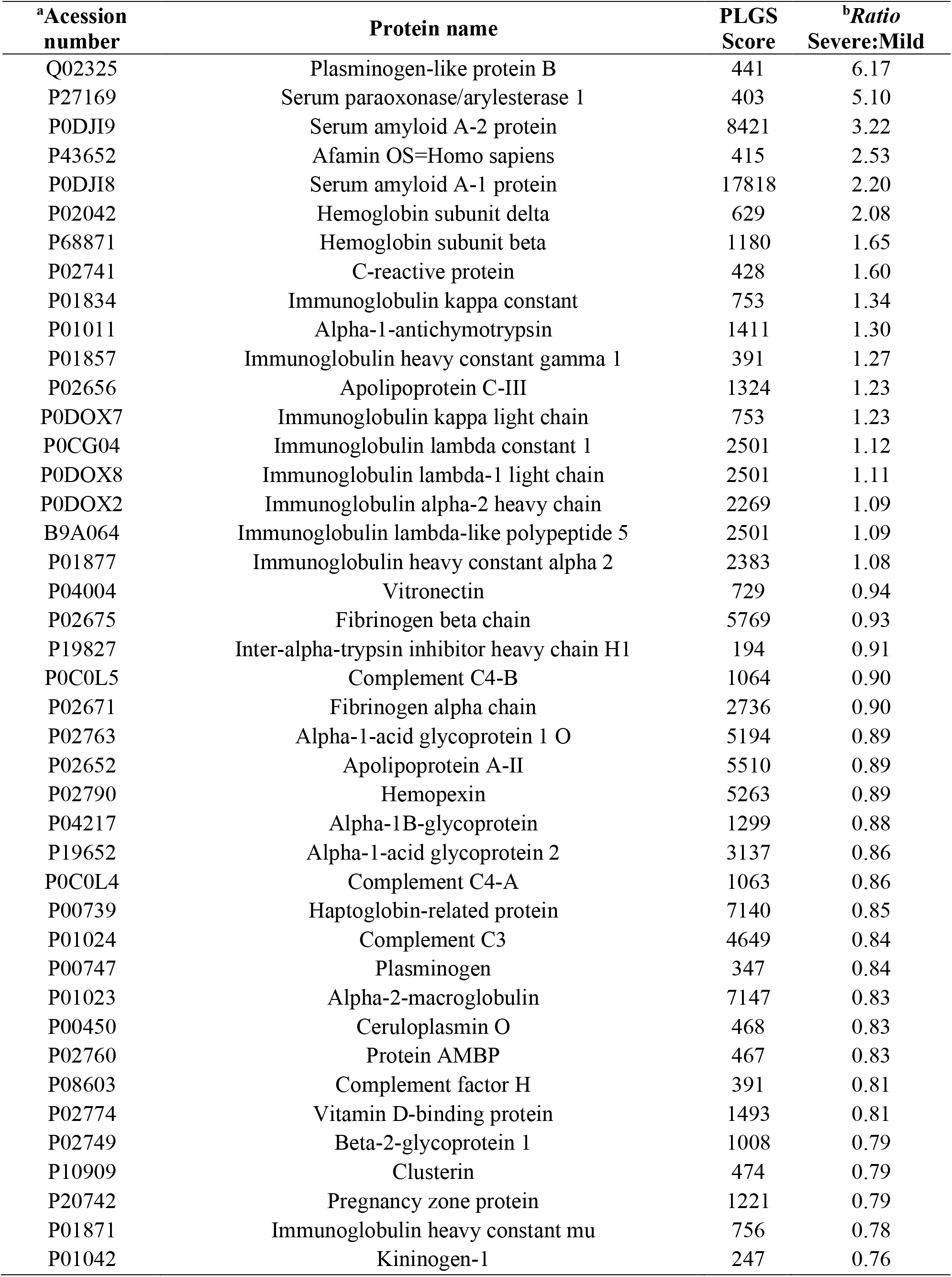
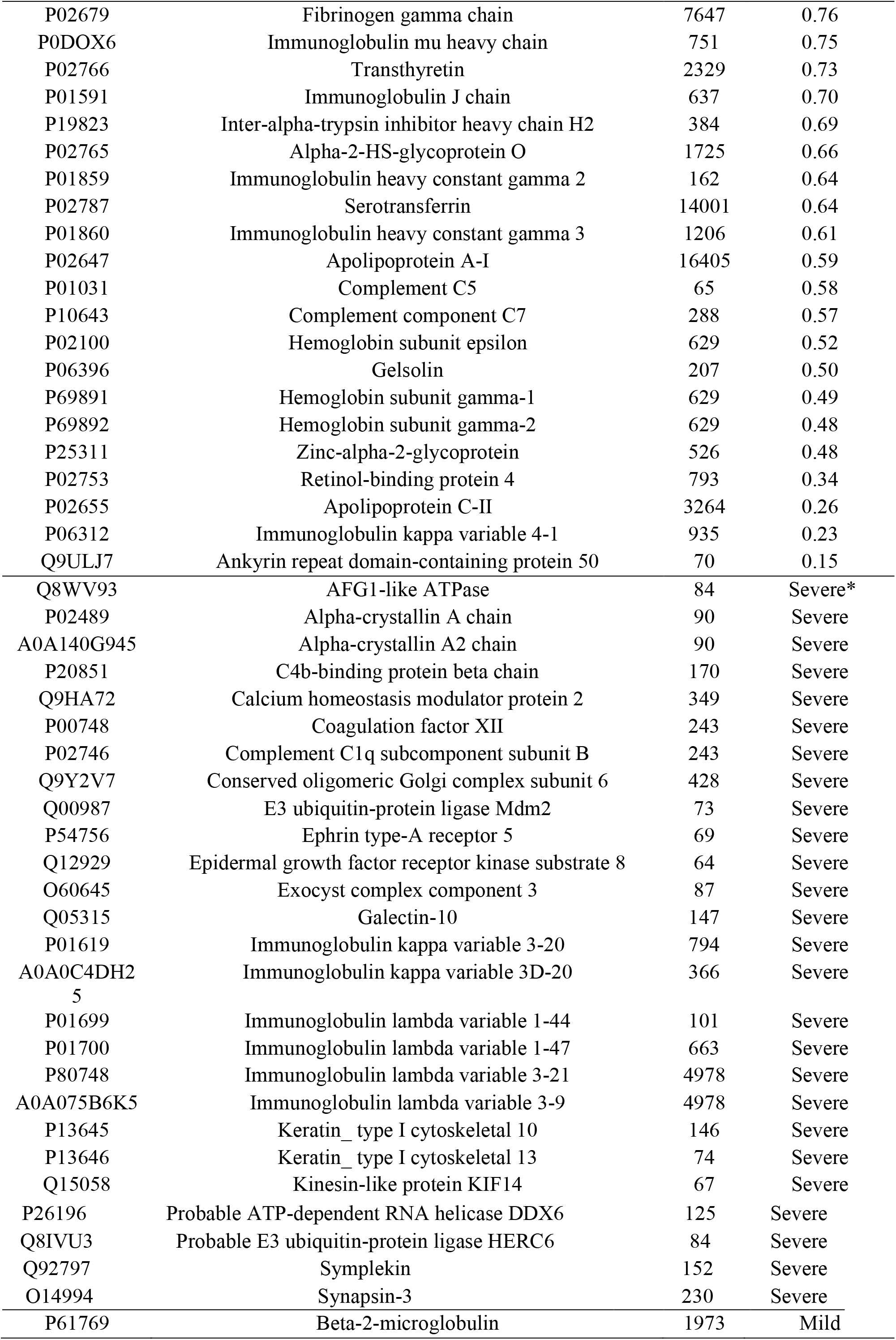
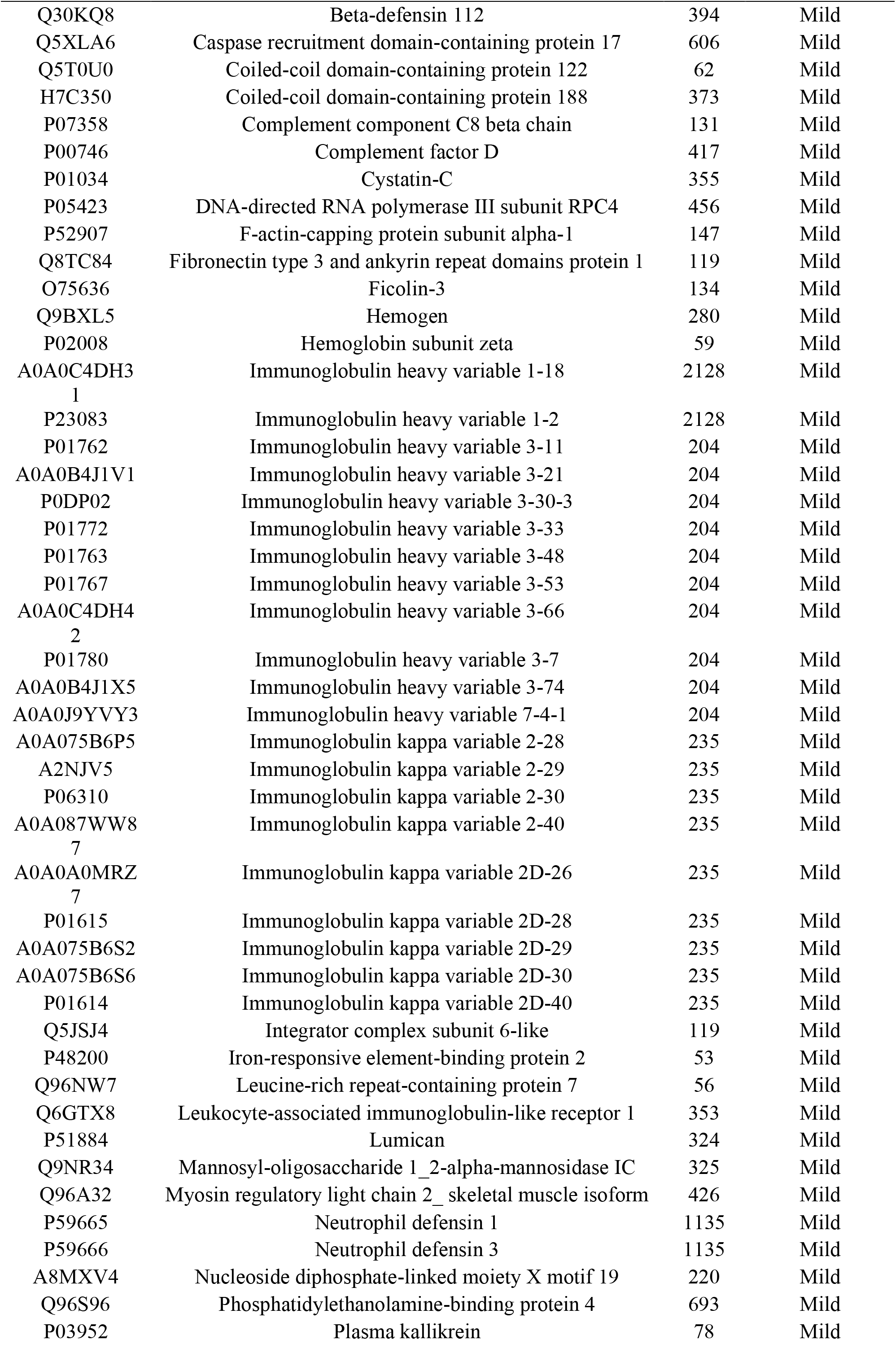
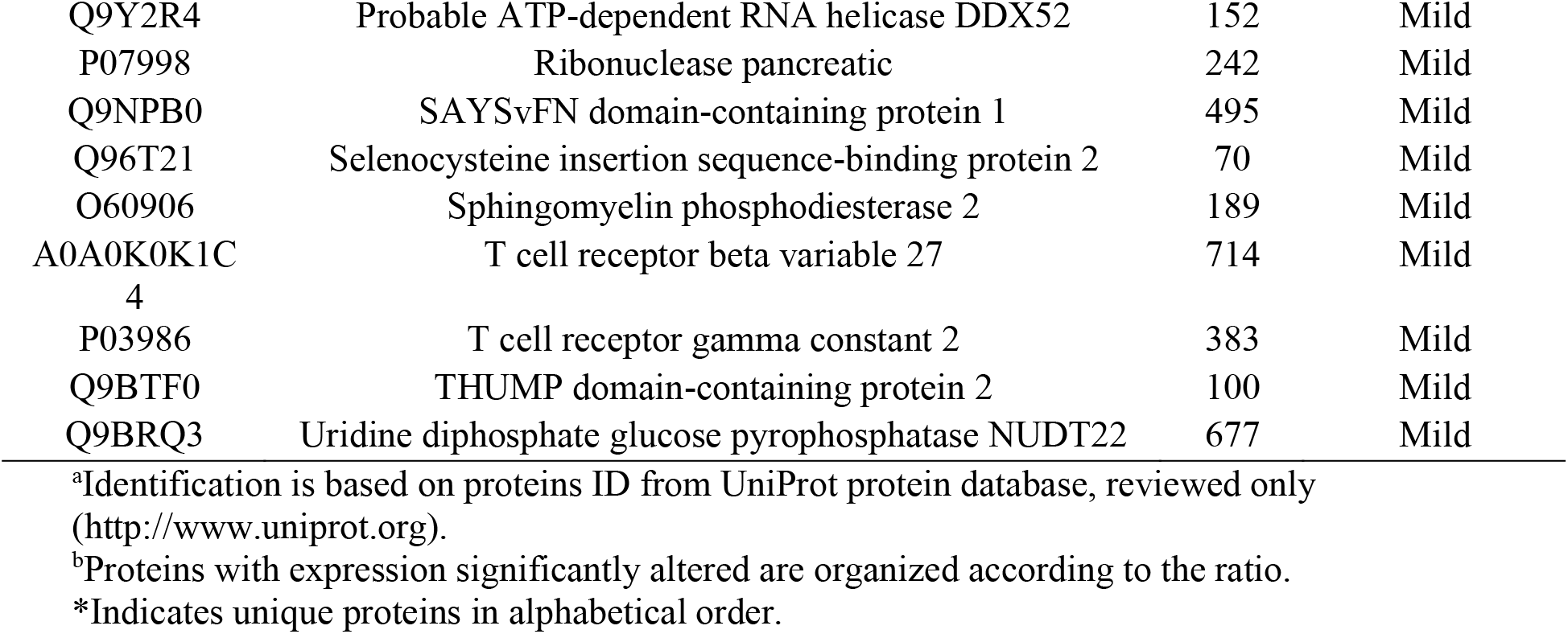
Proteins with expression significantly altered in the plasma of patients upon admission in Bauru State Hospital, Brazil, between May 4^th^ and July 4^th^, 2020. Comparison between patients with severe symptoms that were discharged after admission to an intensive care unit (ICU) and patients with mild symptoms who were discharged without admission to an ICU.

Forty-five proteins 45 were decreased in severe patients compared to mild ones (**Table 3**). Among the proteins with the highest decreases are Ankyrin repeat domain-containing protein 50 (ANKRD50; Q9ULJ7, 6.7-fold), Apoliprotein C-II (P02655; 3.8-fold), Retinol-binding protein 4 (P02753; 2.9-fold) and Gelsolin (GELS; P06396; 2-fold). Proteins decreased but with lower fold-changes are mainly related to phagocytosis [Inter-alpha-trypsin inhibitor heavy chain H1 (P19827), Alpha-2-HS-glycoprotein (P02765)], immune system [Vitronectin (P04004), several components of the complement system, Protein AMBP (P02760), Alpha-1B-glycoprotein (P04217), Zinc-alpha-2-glycoprotein (P25311)], blood coagulation [several isoforms of fibrinogen, Plasminogen (P00747), Kininogen-1 (P01042), Beta-2-glycoprotein (P02749), Clusterin (P10909)], endopeptidase inhibitor activity [Pregnancy zone protein (P20742), Alpha-2-macroglobulin (∝_2_-M; P01023)] and acute phase inflammatory response (Alpha-1-acid glycoprotein 1 (AGP-1; P02763) and 2 (AGP-2; P19652) and Transthyretin (P02766)]. Serotransferrin (P02787) was also decreased. Moreover, 55 proteins were only identified in the plasma collected from patients with mild symptoms (Table 3). Among them are those related to antimicrobial humoral response [Beta-2-microblobulin (P61769), Leukocyte-associated immunoglobulin-like receptor 1 (Q6GTX8), Neutrophil defensin 1 (P59665), Neutrophil defensin 2 (P59666)], innate immune response [Beta-defensin 112 (Q30KQ8), Complement component C8 beta chain (P07358), Complement factor D (P00746), DNA-directed RNA polymerase III subunit RPC4 (POLR3D; P05423), F-actin-capping protein subunit alpha-1 (CAZA1; P52907), Ficolin-3 (FCN3; O75636)], adaptive immune response [T cell receptor beta variable 27 (A0A0K0K1C4) and T cell receptor gamma constant 2 (P03986)], endopeptidase inhibitor activity [Caspase recruitment domain-containing protein 17 (Q5XLA6), Cystatin-C (P01034], iron metabolism [Iron-responsive element-binding protein 2 (IREB2; P48200)], neutrophil degranulation [Leucine-rich repeat-containing protein 7 (Q96NW7)] and blood coagulation [Plasma kallikrein (P03952)] (**Table 3**). In the interaction subnetwork, proteins with change in expression interacted mainly with Microtubule-associated protein tau (P10636), Nuclear factor NF-kappa-B p105 subunit (P19838) and Histone-lysine N-methyltransferase NSD2 (O96028) (**Figure 5**).

**Figure 5.**
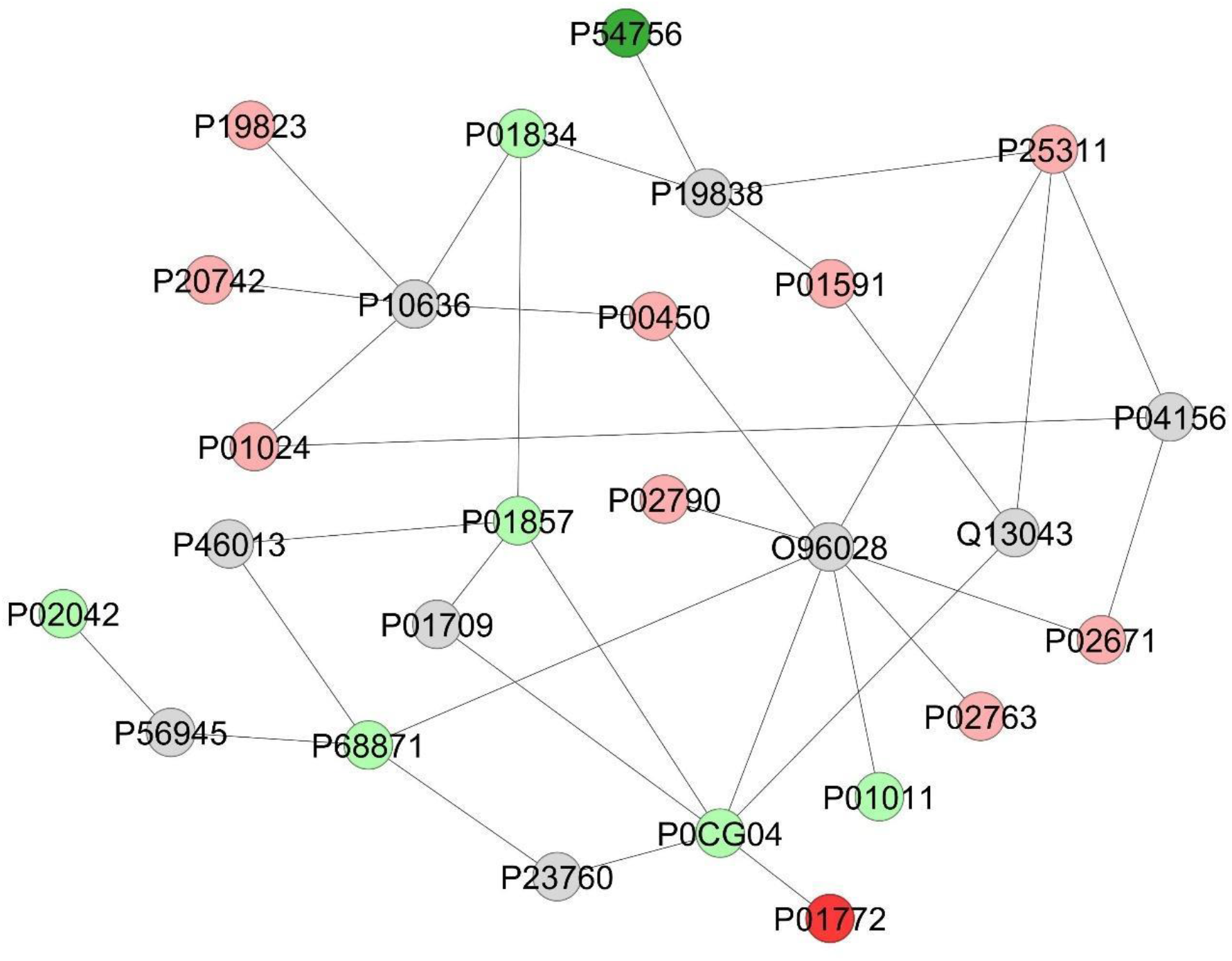
Subnetwork created by ClusterMarker to establish the relationship among proteins identified with differential expression in the plasma patients admitted to Bauru State Hospital, Brazil, between May 4^th^ and July 4^th^, 2020, who were diagnosed with severe or mild COVID-19. The color of the nodes indicates difference in expression of the respective protein defined by its access code (UNIPROT ID). The dark red and dark green nodes indicate proteins unique to mild and severe groups, respectively. Light red and light green nodes indicate down and upregulation in the severe group in respect to mild. The grey nodes indicate interacting proteins that were offered by CYTOSCAPE but were not identified in the present study.

As for the comparison between critical *vs*. mild patients (**Table 4**), 46 proteins were increased and 8 proteins were exclusively found in the first. The proteins with the highest increases were Beta-2-microglobulin (P61769; 6.9-fold), Complement C1s subcomponent (P09871; 3.1-fold), Serum amyloid A-1 (P0DJI8; 2.7-fold) and Serum amyloid A-2 (P0DJI9; 2.2-fold). Other increased proteins were Retinol-binding protein 4 (P02753), Vitamin D-binding protein (VDBP; P02774), Inter-alpha-trypsin inhibitor heavy chain H1 (ITIH1; P19827) and H2 (ITIH2; P19823), Alpha-1B-glycoprotein (A1BG; P04217), proteins involved in coagulation, such as Fibrinogen alpha, beta and gamma chains (P02671, P02679 and P02675, respectively), Kininogen-1 (P01042), Plasminogen (P00747), Phosphatidylethanolamine-binding protein 4 (PEBP4; Q96S96) and Beta-2-glycoprotein (P02749), as well as several components of the complement system, such as Complement C4-A (P0C0L4), Complement C4-B (P0C0L5), Complement Factor B (P00751) and Complement Factor H (P08603). Endopeptidase inhibitors, such as Pregnancy zone protein (P20742), ∝_2_-M (P01023) and Alpha-1-antitrypsin (∝_1_-AT; P01009) were also increased. Among the proteins uniquely found in the critical patients are Gal-10 (Q05315), as well as several proteins related to cardiovascular disease, such as Myoglobin (P02144), Asialoglycoprotein receptor 2 (ASGR2; P07307), Coatomer subunit epsilon (COPE; O14579) and V-set and transmembrane domain-containing protein 4 (VSTM4; Q8IW00).

**Table 4.**
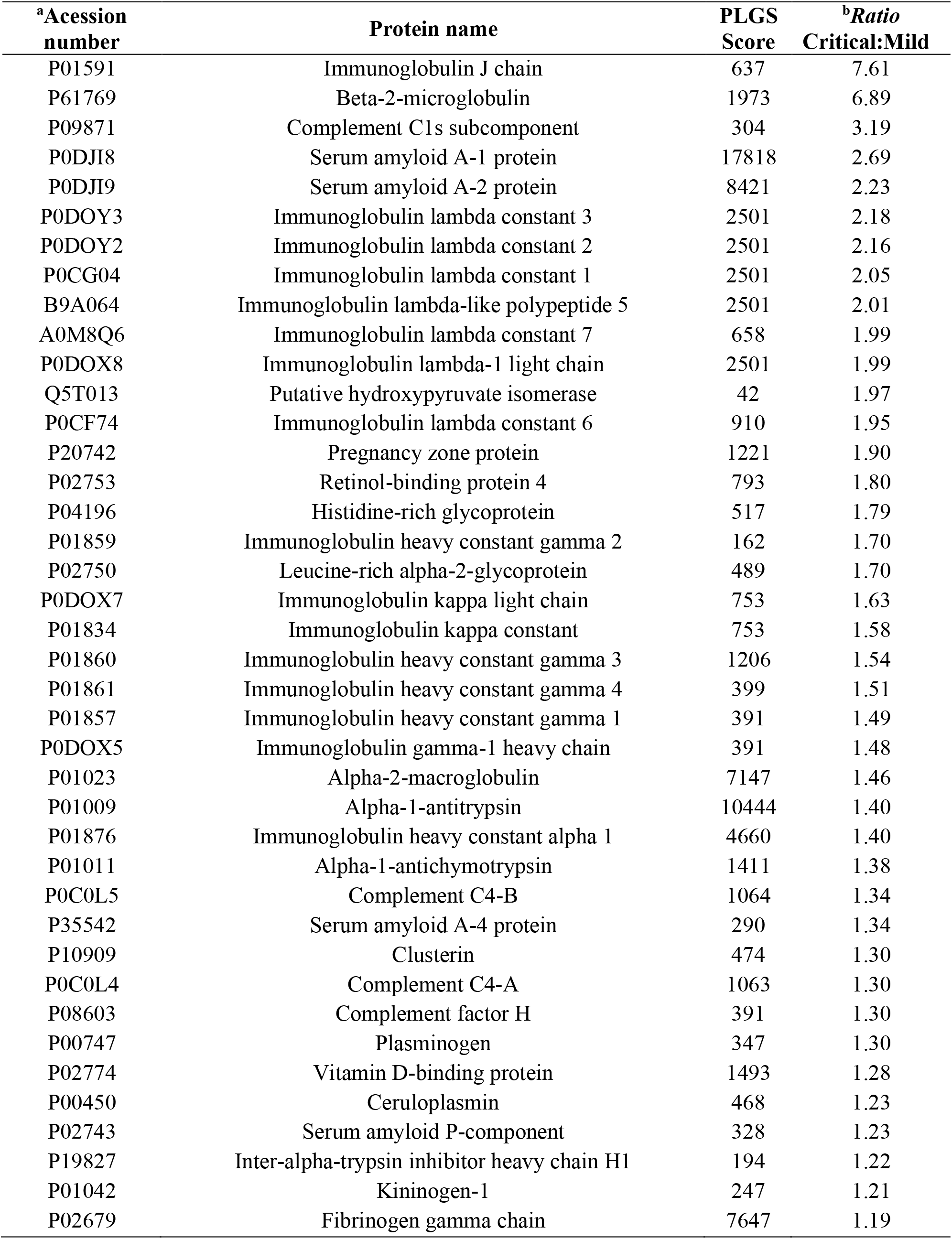
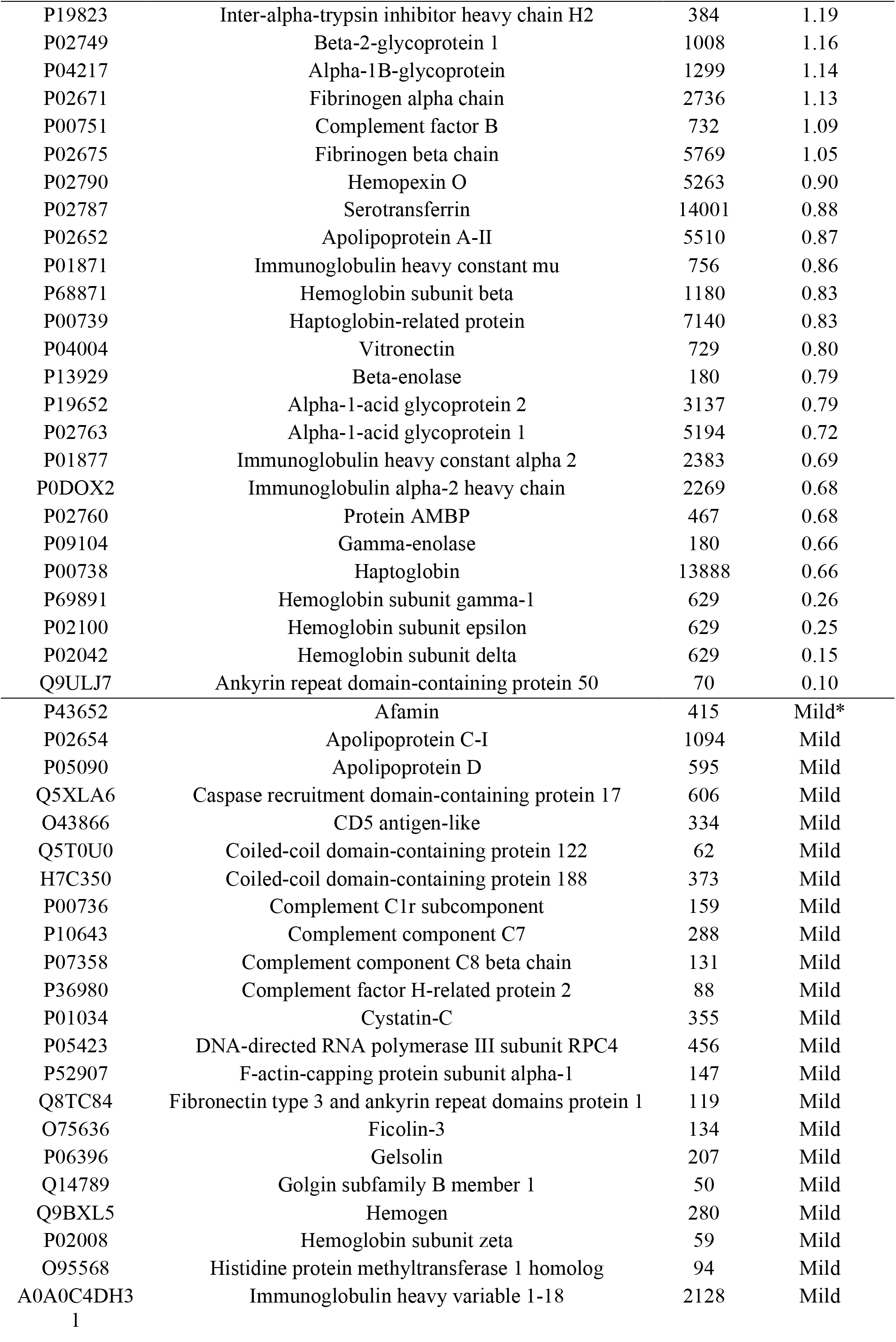
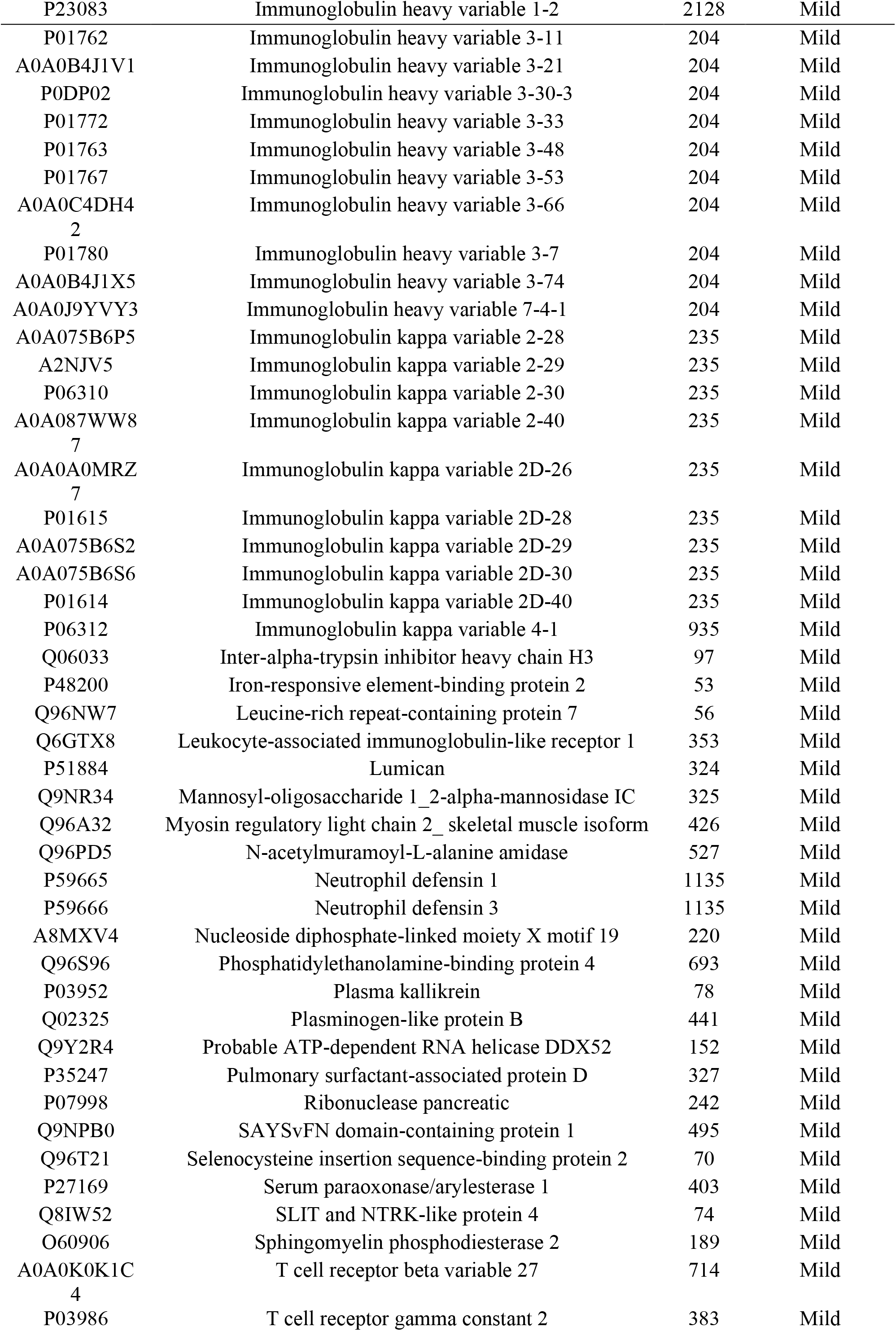
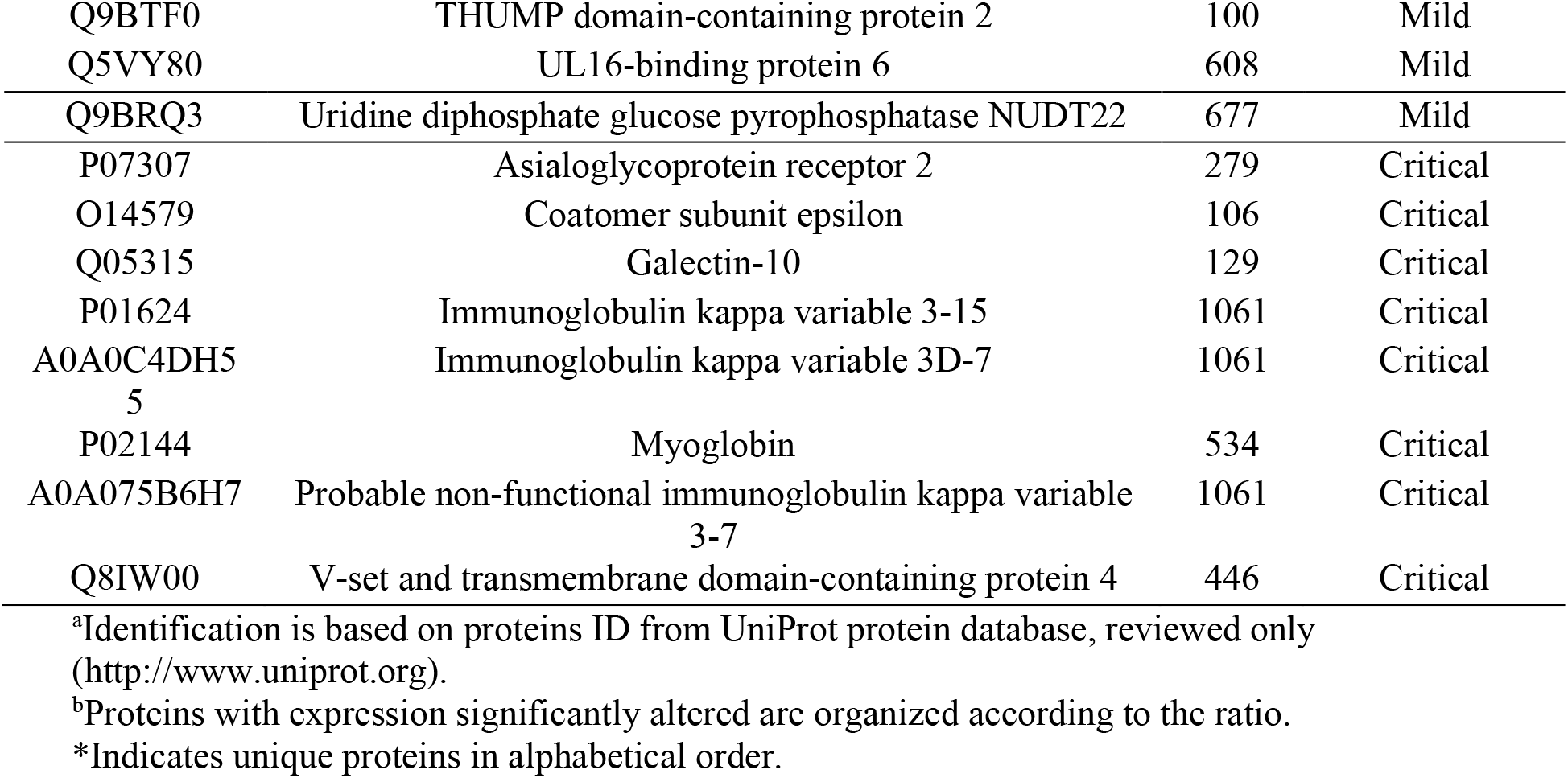
Proteins with expression significantly altered in the plasma of patients upon admission to Bauru State Hospital, Brazil, between May 4^th^ and July 4^th^, 2020. Comparison between critical patients who were admitted to an intensive care unit (ICU) and died and patients with mild symptoms who were discharged without admission to an ICU.

On the other hand, 19 proteins were reduced in the critical patients compared to those with mild symptoms, while 71 proteins were only found in the latter (**Table 4**). The proteins with the highest decreases were ANKRD50 (Q9ULJ7, 10-fold) and subunits of hemoglobin delta, epsilon and gamma, with fold changes ranging between 3 and 6. Other decreased proteins were Haptoglobin (P00738), Beta-enolase (P13929), Gamma-enolase (P09104), Alpha-1-acid glycoprotein 1 (AGP-1; P02763) and 2 (AGP-2; P19652), Protein AMBP (P02760), Vitronectin (P04004), Apolipoprotein A-II (P02652) and Serotransferrin (P02787). Most of the proteins exclusively found in the mild patients are those reported as unique proteins in the same group in the previous comparison (severe *vs*. mild patients). In addition to these, other unique proteins found in mild patients only in this comparison were Apolipoprotein C-I (P02654), Apolipoprotein D (P05090), CD5 antigen-like (CD5L; O43866), GELS (P06396), N-acetylmuramoyl-L-alanine amidase (PGRP2; Q96PD5), Plasminogen-like protein B (PLGB; Q15195), Pulmonary surfactant-associated protein D (SFTPD; P35247), Serum paraoxonase/arylesterase 1 (PON1; P27169) and UL16-binding protein 6 (ULBP6; Q5VY80) (**Table 4**). In the interaction subnetwork, proteins with change in expression interacted mainly with Microtubule-associated protein tau (P10636), Estrogen receptor (P03372), Glycogen synthase kinase-3 beta (P49841), Protein MS12homolog (Q9H081) and A-kinase anchor protein 1, mitochondrial (Q92667) (**Figure 6**).

**Figure 6.**
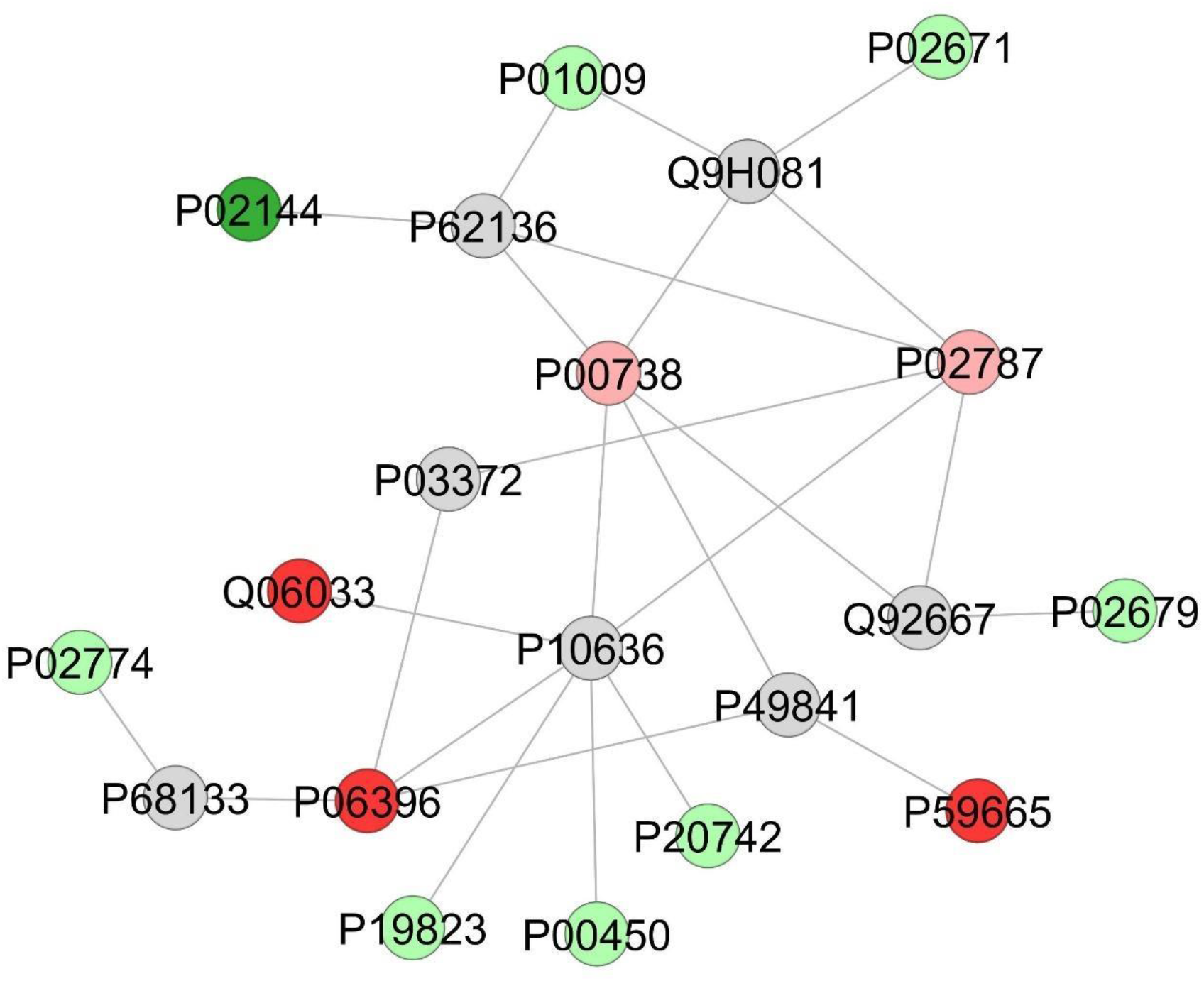
Subnetwork created by ClusterMarker to establish the relationship among proteins identified with differential expression in the plasma patients admitted to Bauru State Hospital, Brazil, between May 4^th^ and July 4^th^, 2020, who were diagnosed with critical or mild COVID-19. The color of the nodes indicates difference in expression of the respective protein defined by its access code (UNIPROT ID). The dark red and dark green nodes indicate proteins unique to mild and critical groups, respectively. Light red and light green nodes indicate down and upregulation in the critical group in respect to mild. The grey nodes indicate interacting proteins that were offered by CYTOSCAPE but were not identified in the present study.

When critical patients were compared to severe ones, 47 proteins were increased and 10 proteins were exclusively found in the first. The proteins with the highest increases were associated with blood coagulation, such as Heparin cofactor 2 (P05546; 5-6-fold) and Plasminogen-like protein A (Q15195, 3.6-fold) as well as with humoral immune response, such as Retinol-binding protein 4 (P02753; 5.3-fold). Other increased proteins were also increased in the critical patients when compared to mild ones. Among them are VDBP, ITIH2, A1BG, proteins involved in coagulation, such as Fibrinogen alpha, beta and gamma chains (P02671, P02679 and P02675, respectively), as well as several components of the complement system, such as Complement C4-A (P0C0L4), Complement C4-B (P0C0L5) and Complement Factor B (P00751). Endopeptidase inhibitors, such as Pregnancy zone protein (P20742), ∝_2_-M and Alpha-1-antitrypsin (∝_1_-AT; P01009) were also increased. Other increased proteins in the critical patients when compared to the severe ones, not found in the comparison between critical *vs*. mild patients were Angiotensinogen (P01019), proteins related to phagocytosis [Alpha-2-HS-glycoprotein (P02765)], Apolipoproteins A-I (P02457), A-IV (P06727), C-II (P02655) and E (P02649), proteins related to blood coagulation [Clusterin (P10909), Fibronectin (P02751), Histidine-rich glycoprotein (P04196), Plasma protease C1 inhibitor (P05155)], several components of the complement system [Complement C1s subcomponent (P09871, Complement C3 (P01024), Complement C5 (P10131) and Complement component C9 (P02748)] and proteins involved in acute inflammatory response [Transthyretin (P02766)]. Among the proteins exclusively found in the critical patients when compared to the severe ones are the same ones found only in the critical patients in comparison to the mild patients, except for Gal-10. In addition, Beta-2-microglobulin (P61769), Beta-defensin 112 (Q30KQ8), Complement factor D (P00746) were also exclusively found in critical patients in this comparison (**Table 5**).

**Table 5.**
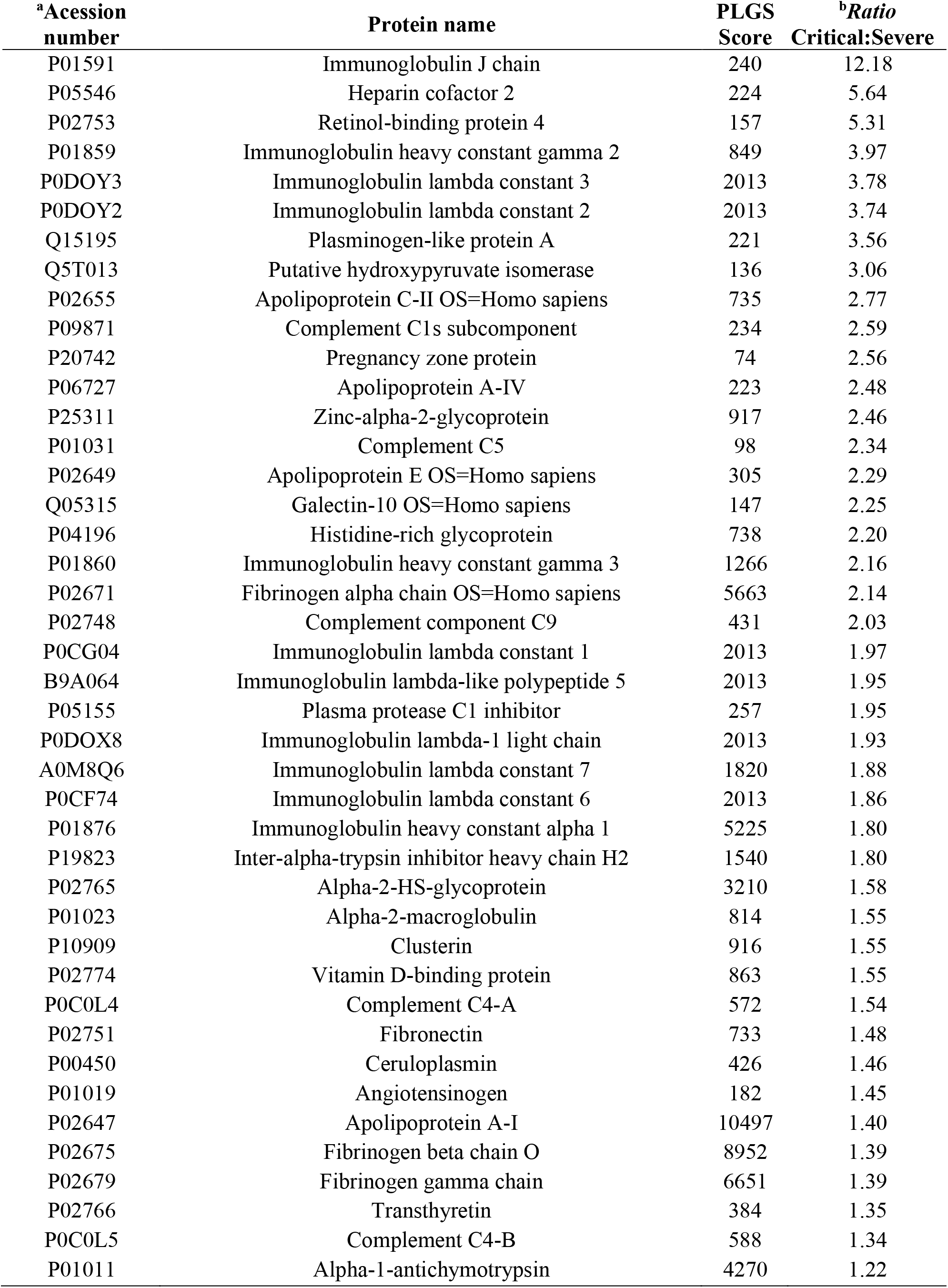
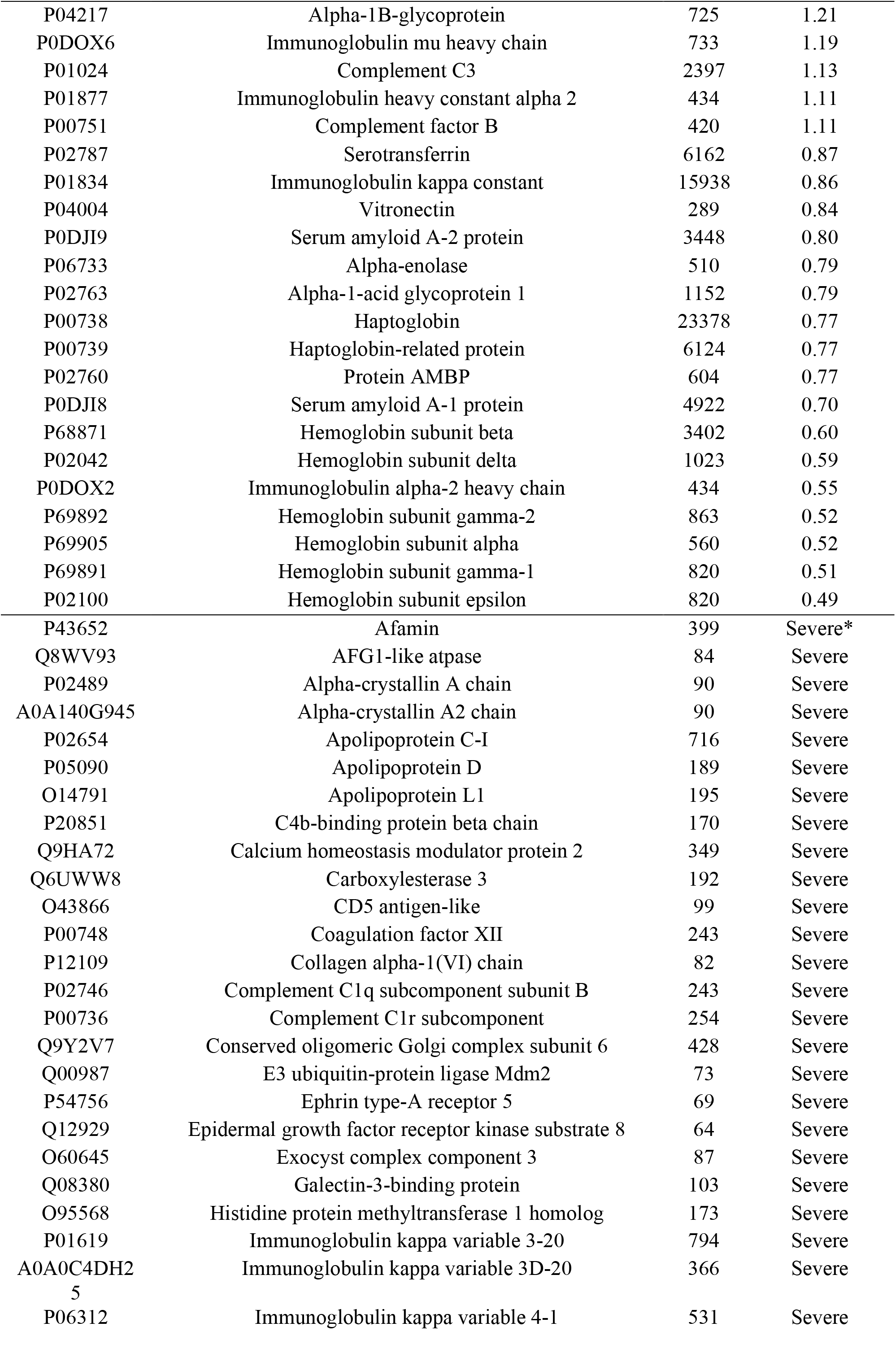
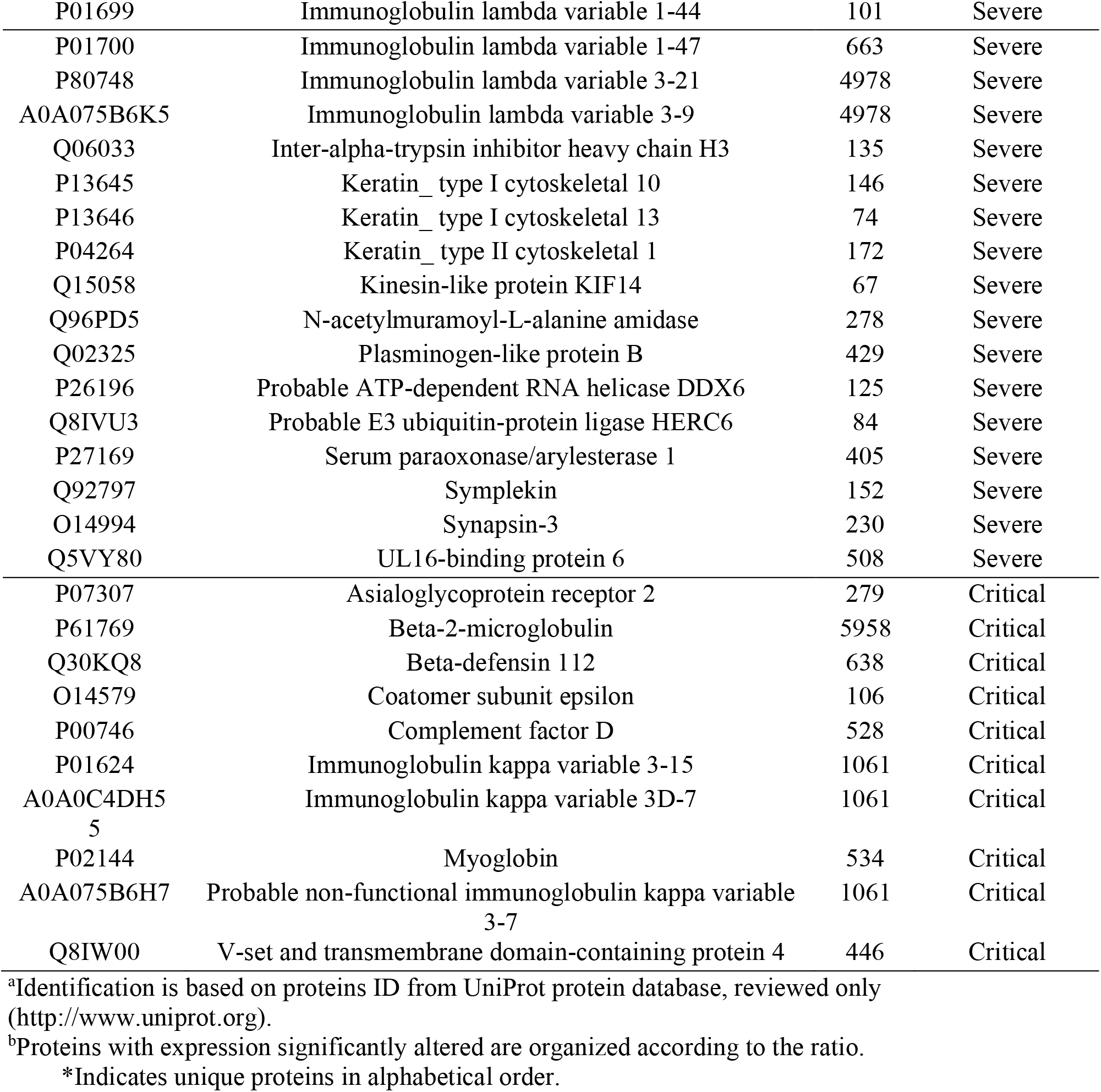
Proteins with expression significantly altered in the plasma of patients upon admission to Bauru State Hospital, Brazil, between May 4^th^ and July 4^th^, 2020. Comparison between critical patients who were admitted to an intensive care unit (ICU) and died and patients with severe symptoms that were discharged after admission to an ICU.

On the other hand, 17 proteins were decreased in critical patients, compared to severe ones, and 42 proteins were exclusively found in the latter. Among the decreased proteins are several subunits of hemoglobin, alpha-enolase (P06733), Serotransferrin (P02787), acute inflammatory response proteins, such as Serum amyloid A-1 (P0DJI8) and A-2 (P0DJI9) and AGP1 (P02763), as well as proteins with immunomodulatory effects, such as Vitronectin (P04004) and AMBP (P02760). Among the proteins exclusively found in severe patients when compared to critical ones, are CD5 antigen-like (CD5L; O43866), apolipoproteins C-I (P02654), D (P05090) and L1 ()14791), proteins related to blood coagulation [C4b-binding protein beta chain (P20851), Coagulation factor XII (P00748), PLGB (Q15195)], complement activation [Complement C1q subcomponent subunit B (P02746)], regulators of endopeptidase activity [Complement C1r subcomponent (P00736), ITIH3 (Q06033)] and immune response [PGRP2, (Q96PD5) and ULBP6 (Q5VY80) (**Table 5**). In the interaction subnetwork, proteins with change in expression interacted mainly with Microtubule-associated protein tau (P10636), Estrogen receptor (P03372), PAS domain-containing serine/threonine-protein kinase (Q96RG2), Nuclear factor of kappa light polypeptide gene enhancer in B-cells 2 (P49/p100), isoform CRA_c (D3DR86), Microtubule-associated proteins 1A/1B light chain 3A (Q9H492), Gamma-aminobutyric acid receptor-associated protein-like 1 (Q9H0R8), N6-adenosine-methyltransferase catalytic subunit (Q86U44), Protein phosphatase 1 regulatory subunit 15A (O75807), Vascular cell adhesion protein 1 (P19320), Albumin (P02768), Histone-lysine N-methyltransferase NSD2 (O96028), Protein MIS12 homolog (Q9H081) and A-kinase anchor protein 1, mitochondrial (Q92667) (**Figure 7**).

**Figure 7.**
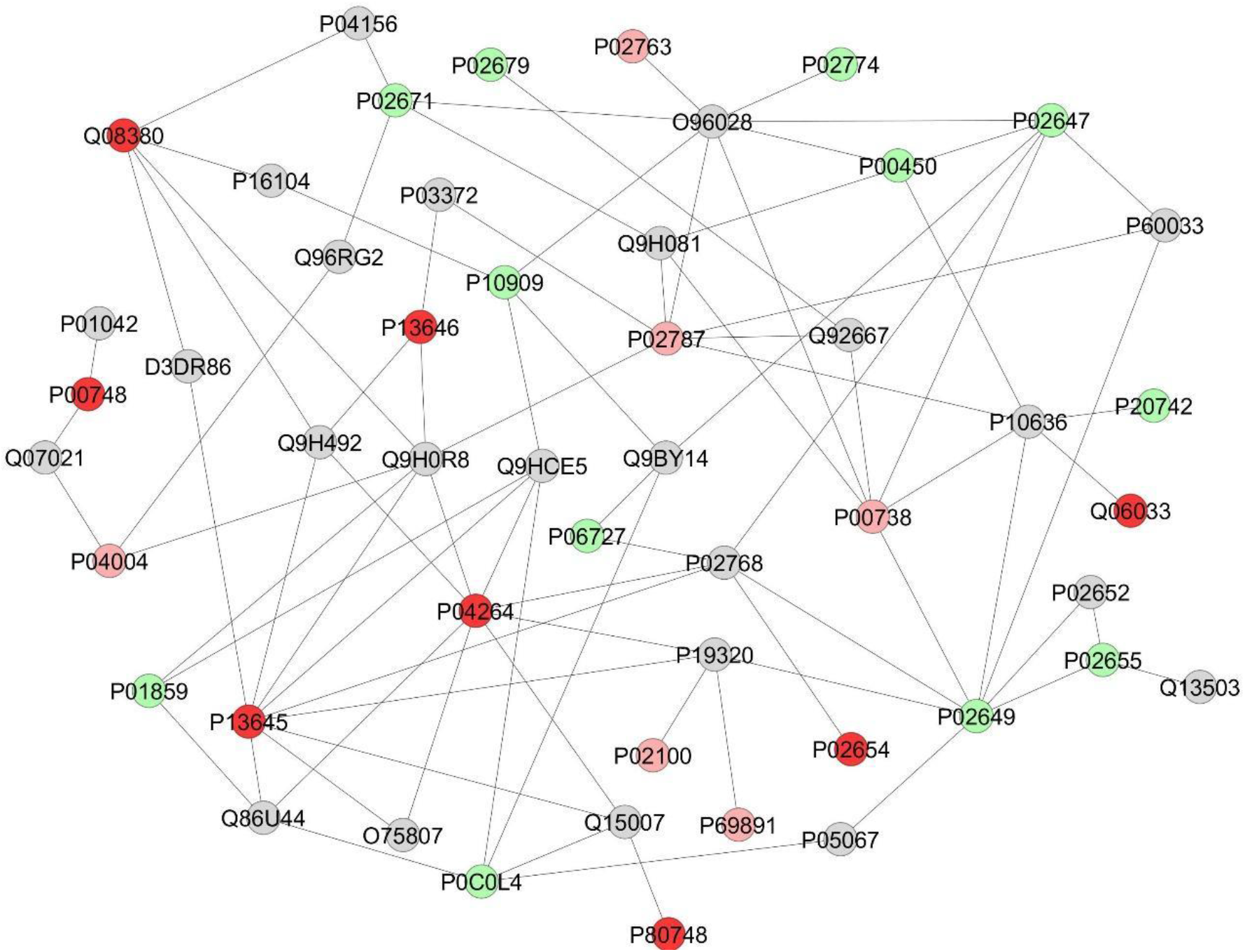
Subnetwork created by ClusterMarker to establish the relationship among proteins identified with differential expression in the plasma patients admitted to Bauru State Hospital, Brazil, between May 4^th^ and July 4^th^, 2020, who were diagnosed with critical or severe COVID-19. The color of the nodes indicates difference in expression of the respective protein defined by its access code (UNIPROT ID). The dark red and dark green nodes indicate proteins unique to severe and critical groups, respectively. Light red and light green nodes indicate down and upregulation in the critical group in respect to severe. The grey nodes indicate interacting proteins that were offered by CYTOSCAPE but were not identified in the present study.

## Discussion and Conclusion

The present study was designed to find out biomarkers of prognosis for COVID-19 patients upon admission to hospital, as well as to search possible therapeutical targets. We had a prospective cohort design. All patients admitted to the HEB within a 2-month period were enrolled. Upon admission, we collected blood samples for laboratory and proteomic analyses and the results were related to the course of the disease, being the patients classified into 3 severity categories: mild (survivors, no need of admission to an ICU), severe (survivors, after admission to an ICU) or critical (non-survivors, after admission to an ICU). Critical patients who died were older than mild and severe ones and there was also a predominance of men, in-line with previous studies (6, 17, 18). The higher susceptibility of males in comparison to females has been attributed to the protective effect of estrogen in women or to stronger immune response with higher levels of cytokines in men (19, 20). The most common comorbidity among the patients was hypertension, followed by diabetes (Table 1), as has been shown in previous studies (6, 17, 18). Alterations in the rennin-angiotensin system (RAS), with consequent activation of NLRP3 (NOD-like receptor 3) inflammasome is the probable reason why hypertensive patients are more susceptible to severe forms of COVID-19 (21).

In the laboratory exams (**Table 2**), the hemoglobin levels did not change among the groups, which is consistent with more recent findings showing no evidence of hemoglobin damage by SARS-CoV-2 infection (22). In the present study, severe and critical patients had increased plasma ferritin levels compared to mild ones, as has been reported in other studies (23, 24). Ferritin is a mediator of immune dysregulation. Increased ferritin levels contribute to the cytokine storm via direct immune-suppressive and pro-inflammatory effects (25). The increased levels of ferritin in severe and critical patients might be due to the lower expression of IREB2. This protein was found exclusively in the plasma of mild patients. It binds to iron-responsive elements (IRES) in the 5’-UTR of ferritin mRNAs, repressing translation (26). Thus, increasing the expression of IREB2 might be a therapeutic possibility to reduce ferritin levels and, in turn, the severity of COVID-19 (**Table 6**). Critical patients had also decrease in serotransferrin when compared to mild ones, in-line with findings showing high ferritin and low transferrin levels associated with increased risk for ICU admission and need for mechanical ventilation (27).

**Table 6.** Main plasma proteomic findings upon admission, implications on the course of COVID-19 and potential associated therapies.

| **Plasma proteomic findings** | **Implications on the course of COVID-19** | **Potential therapies** |
| --- | --- | --- |
| IREB2 was found exclusively in patients with mild symptoms. | This protein binds to iron-responsive elements in the 5'-UTR of ferritin mRNAs, thus repressing translation (26). Its absence in severe and critical patients might be responsible for the higher plasma ferritin levels found in these patients in the laboratory exams. Ferritin is a mediator of immune dysregulation. Increased ferritin levels contribute to the cytokine storm via direct immune-suppressive and pro-inflammatory effects (25). | Increasing the expression of IREB2 might help reducing ferritin levels and, in turn, the severity of COVID-19. |
| GELS was found exclusively in patients with mild symptoms. | Calcium-binding protein that scavenges circulating filamentous actin, thus possessing anti-inflammatory properties. Reduced levels of GELS have been shown in serum (9, 11) and plasma (10) of COVID-19 patients with worse outcomes. | GELS supplementation has been suggested as a potential therapy for COVID-19 (9) and a clinical trial of recombinant plasma form GELS is currently being conducted (NCT04358406). |
| POLR3D was found exclusively in patients with mild symptoms. | This enzyme is involved in innate immune response, playing a key role in limiting infection by intracellular bacteria and viruses. It is involved in the production of type I interferon (26). Its increased expression might be associated with more favorable prognosis of COVID-19. | Increasing the expression of POLR3D in the initial stages of the disease. |
| SFTPD was only identified in mild patients compared to critical ones. | Participates in the turnover of pulmonary surfactant and modulates the action of leukocytes in immune response, contributing to lung's defense against inhaled microorganisms (26). | Increasing the expression of SFTPD. |
| PON1 was only identified in mild patients compared to critical ones. | This enzyme possesses arylalkylphosphatase activity, being involved in the protection of low-density lipoproteins against oxidative damage and the formation of atheroma (26). It is also important for innate immune response (47). In a recent study involving in silico discovery of candidate drugs against COVID-19, it was reported that genes correlated with *ACE2* are enriched in arylalkylphosphatase activity (48). | Increasing the activity of PON1. |
| ULBP6 was only identified in mild patients compared to critical ones. | Is a member of the MHC class I superfamily that binds and activates the natural killer group 2 member D (NKG2D) receptor, localized in the surface of immune cells, mediating natural killer cell cytotoxicity (49). NKG2D ligands play a central role in the immunological control. SNPs within the ULBP6 gene have a profound influence on the clinical outcome of allogeneic stem cell transplantation (50) and are also associated to diabetic nephropathy (51) and alopecia areata (52). Moreover, arginine-to-leucine polymorphism within ULBP0602 increases the affinity of the NKG2D/ULBP6 interaction, thus reducing NKG2D-mediated activation (53). It is possible that critical patients might have this polymorphism leading to increased affinity and, for this reason, ULBP6 was not found in their plasma, thus leading to impaired immunological response to SARS-CoV-2 | Modulating the interaction ULBP6/NKG2D receptor. |
| Gal-10 was identified exclusively in critical and severe patients, compared to the mild ones. It was also increased in critical patients, compared to severe ones, i.e., it showed a dose-response pattern, with increasing levels accompanying the increase in the severity of the disease. | CLC is a hallmark of eosinophil death. It suffers a phase transition to a crystalline state, originating the Charcot-Leyden crystals (CLCs) that promote type 2 immunity. Moreover, CLCs have proinflammatory effects in the lungs, leading to an influx of neutrophils and monocytes after 6 and 24 h, respectively, after injection, which is accompanied by the production of large quantities of IL-6 and TNF-α in bronchoalveolar lavage fluid and of IL-1β and of CCL2 (monocyte chemoattractant) in lung tissue. Furthermore, CLCs boost adaptative immunity to co-administered antigens (57). Thus, the increase in Gal-10 in critical and severe patients, in a dose-response fashion, might be associated to the “cytokine storm” that is characteristic of the poorer outcomes of COVID-19. Recently, a pre-clinical study showed that antibodies against Gal-10 to preformed CLCs completely dissolved the crystals present in the mucus within 2 hours (57). | Antibodies against Gal-10 should be evaluated as a potential therapy for SARS-CoV-2 positive patients that are diagnosed with high levels of CLCs at admission. |

One of the proteins that presented the highest increases in critical patients compared to those presenting mild symptoms (**Table 4**) and that was exclusively found in critical patients compared to severe ones (**Table 5**) was Beta-2-microglobulin. This protein is a component of the class I major histocompatibility complex (MHC), involved in the presentation of peptide antigens to the immune system (26). It is regarded as a biomarker of the immune system activation in HIV (28, 29) and more severe COVID-19 patients, in which it is increased in the cerebrospinal fluid (28). Its increase in critical patients might be due to the ability of SARS-CoV-2 to evade the immune system in these patients. Our immune system comprises both innate and acquired immune response. The first makes infected cells secrete interferons (IFNs) and pro-inflammatory cytokines. IFNs make non-infected cells develop an antiviral stage, while proinflammatory cytokines activate macrophages and other phagocytes to remove virus and infected cells. CD4+ Th lymphocytes (adaptive responses) are also activated by cytokines and, in turn, activate B-lymphocytes that will produce neutralizing antibodies to fight the virus and CD8+Tc cells to start programmed cell death of cells infected by virus. In other words, innate response exposes the virus and organizes the acquired response to fight the infection. However, the outcome depends on a timely coordination between both immune responses. High production of IFNs in 18-24 h post infection leads to effective innate and acquired immune responses. On the other hand, a delay in the production of IFNs (3-4 days post infection) results in ineffective innate and acquired immune responses (30), despite persistent IL-6 and TNF-α release by several cells, infiltrating monocytes, inflammatory reactions and a dysfunctional response amplified by macrophages (31), which may lead to the critical form of COVID-19. In this case, there is a huge increase in cytokines release, which is known as cytokines release syndrome (CRS), or “cytokines storm”. In the initial stages of inflammation, IL-6 expression is restricted. However, in later stages it is also expressed in the liver, where it elicits the production of acute phase proteins, such as fibrinogen and serum amyloid A (SAA) (30) that were also increased in the present study in critical patients, compared to mild ones (**Table 4**). SAA was also increased in severe patients when compared to mild ones (**Table 3**). These findings agree with previous studies that indicated SAA as a predictor of COVID-19 severity (32, 33). Another classical acute phase protein, PCR, was significantly increased in severe patients compared to mild ones, both in the proteomic and laboratorial analyses (**Tables 3** and **2**, respectively). Moreover, PCR was increased in critical patients compared to mild ones, both in proteomic (data not shown) and laboratorial analyses (**Table 2**), but the difference did not reach statistical significance. The laboratorial analysis validates the proteomic analysis for this protein. Other increased proteins associated to immune response in patients who died (critical) in comparison to survivors not admitted in an ICU (mild) and also to survivors admitted in an ICU (severe) were VDBP (enhancement of the chemotactic activity of C5 alpha for neutrophils in inflammation and macrophage activation) (26), several members of the complement system, as well as A1BG that plays a role in the degranulation of neutrophils and platelets (26). Moreover, POLR3D was only found in patients with mild symptoms, when compared both to severe and critical ones. This enzyme is involved in innate immune response, playing a key role in limiting infection by intracellular bacteria and viruses. It is involved in the production of type I interferon (26). Thus, increased expression of this enzyme might be associated with a more favorable prognostic of COVID-19. Interestingly, when critical patients were compared to mild ones, CD5L, a secreted protein that acts as a key regulator of lipid synthesis, was found exclusively in the latter (**Table 4**). For the comparison critical *vs*. severe patients, CD5L was found only in the latter as well (**Table 5**). This protein is expressed by macrophages mainly in inflamed tissues and regulates mechanisms in inflammatory responses, as occurs in infections (26). Furthermore, patients who died had decreases in proteins with important immunomodulatory activities, such as AGP1 and AGP2 (34), AMBP and vitronectin, when compared to patients with mild symptoms.

The role of ∝_2_-M in COVID-19 has been proposed based on its versatility, i.e., in its tetrameric form it is able to inhibit all four classes of proteases, while in its dimeric form it shows increased interaction with mediators of inflammation, such as TNF-∝, Il-2 and IL-6. Moreover, in children, increased levels of ∝_2_-M are speculated to contribute to the antithrombin activity of plasma and protection against COVID-19, but more studies are necessary to confirm this hypothesis (35). In the present study, however, ∝_2_-M levels were increased in the more severe cases of COVID-19 in all 3 comparisons, which suggests its role in inflammation. Another important role for ∝_2_-M, together with ∝_1_-AT, also increased in critical patients compared to severe and to mild ones (**Tables 4** and **5**), is to control neutrophil elastase, a key enzyme involved in the formation of NETs (neutrophil extracellular traps) (36). Extensive formation of NETs, web-like protease- and histone-coated DNA structures that constitute an immune mechanism to trap pathogens, is found in severe and critical cases of COVID-19 (37), since they can cause platelet activation and thrombosis (38).

Hypercoagulability, with predominance of thrombosis in venous or arterial macro and microcirculation, worsens prognosis of COVID-19 (38-40). The etiology of COVID-19-associated coagulopathy seems to follow Virchow’s Triad, including abnormalities of blood flow, vascular injury and abnormalities within the circulating blood (38). Besides the formation of NETs, other mechanisms are potentially involved in the development of this systemic coagulopathy. Complement-mediated microvascular injury involving lung and skin, with marked deposition of C5b-9, C4d and Mannan-binding lectin serine protease (MASP)-2, suggesting activation of lectin-based and alternative pathways was reported among autopsy findings from decedents with severed COVID-19 and ARDS (41). In the present study, several components of the complement system, as well as their activators were increased in critical patients, compared with those presenting mild symptoms (**Table 4**), such as Complement C1s subcomponent, Complement C4-A, Complement C4-B, Complement Factor B, Complement Factor H and Plasminogen (activates C1 and C5). Another potential mechanism involved in hypercoagulability is endothelial injury provoked by SARS-CoV-2 binding, membrane fusion and viral entry since endothelial cells are characterized by broad expression of ACE2 (angiotensin converting enzyme 2) receptors. This leads to accumulation of inflammatory cells and reticular inclusions (phospholipids and glycoproteins originated from rough endoplasmic reticulum that accumulate in endothelial cells in response to high levels of interferon) that damage endothelial cells and provoke expression of prothrombotic genes (38). In sites of endothelial injury, release of VDBP (increased in critical patients compared to severe and to mild ones) enhances the chemotactic effect of C5a, leading to attraction, aggregation and activation of monocytes and neutrophils, generating an oxidative burst (42). Vitamin D competes for the same binding site on VDBP, reducing this chemotactic effect and controlling the course of the disease (43). In the present study, D-dimer levels were significantly increased in plasma of severe and critical patients, compared to mild ones (**Table 2**), indicating a state of hypercoagulability in the first two groups. This laboratorial result is consistent with the proteomic findings, since D-dimer is a fibrin degradation product found in the peripheral blood, after fibrin is formed from fibrinogen and degraded by plasminogen activators (38).

GELS and CAZA1 were exclusively found in mild patients, compared to severe and critical. These calcium-binding proteins scavenge circulating filamentous actin, thus possessing anti-inflammatory properties. For this reason, reduced levels of GELS have been shown in serum (9, 11) and plasma (10) of COVID-19 patients with worse outcomes, in-line with our results. In fact, GELS supplementation has been suggested as a potential therapy for COVID-19 (9) and a clinical trial of recombinant plasma form GELS is currently being conducted (NCT04358406) (**Table 6**).

Some protective proteins that would be involved in the control of the immune response or blood coagulation were increased, or exclusively identified in patients with mild symptoms, compared to severe (**Table 3**) or critical patients (**Table 4**). These proteins could potentially help to explain why the disease took a milder course in these patients. Among the players involved with the immune response are PGRP2 that digests biologically active peptidoglycan (PGN) into biologically inactive fragments (44) and FCN3, which functions as an opsonin, making easier bacterial elimination via the complement cascade (45). Among the proteins that prevent the formation of clots we found PEBP4, a serine protease inhibitor that inhibits thrombin (46), and Beta-2-glycoprotein that prevents the activation of the intrinsic blood coagulation cascade by binding to phospholipids on the surface of damaged cells (26).

A couple of interesting protective proteins were uniquely found in mild patients compared to critical ones (**Table 4**). One of them is SFTPD that participates in the turnover of pulmonary surfactant and modulates the action of leukocytes in immune response, contributing to lung’s defense against inhaled microorganisms (26). Another protein, PON1, possesses aryadialkylphosphatase activity, being involved in the protection of low-density lipoproteins against oxidative damage and the subsequent series of events leading to the formation of atheroma (26). It is also important for innate immune response and is reduced during hepatitis virus B infection, correlating with the functional status of the liver (47). Moreover, in a recent study involving in silico discovery of candidate drugs against COVID-19, it was reported that genes correlated with *ACE2* are enriched in aryadialkylphosphatase activity (48). ULBP6 is a member of the MHC class I superfamily that binds and activates the natural killer group 2 member D (NKG2D) receptor, localized in the surface of immune cells, mediating natural killer cell cytotoxicity (49). NKG2D ligands play a central role in immunological control. SNPs within the ULBP6 gene have a profound influence on the clinical outcome of allogeneic stem cell transplantation (50) and are also associated to diabetic nephropathy (51) and alopecia areata (52). Moreover, arginine-to-leucine polymorphism within ULBP0602 increases the affinity of the NGK2D/ULBP6 interaction, thus reducing NKG2D-mediated activation (53). It is possible that critical patients might have this polymorphism leading to increased affinity and, for this reason, ULBP6 was not found in their plasma, thus leading to impaired immunological response to SARS-CoV-2 (**Table 6**). Future studies should be conducted to evaluate the occurrence of this polymorphism in patients with critical and mild symptoms of COVID-19 to confirm this hypothesis.

On the other hand, some proteins were only present in critical patients compared to mild ones (**Table 4**) and their trend to increased expression might be associated with the poorer clinical outcome in the first. Among them are several proteins related to cardiovascular disease. One of them is myoglobin, an early biochemical marker of acute myocardial infarction, although non-specific since it is present also in skeletal muscle (54). Its presence in the blood of critical patients might indicate damage to cardiac or skeletal muscles. Other proteins in this category are COPE, increased in patients with atherosclerosis (55) and ASGR2, whose deficiency reduces non-HDL cholesterol and activates platelets (56).

A remarkably interesting finding of the present study was the identification of Gal-10 exclusively in the critical and severe patients, compared to the mild ones (**Tables 4** and **3**, respectively). This protein was also increased in critical patients, compared to severe ones (**Table 5**), i.e., it showed a dose-response pattern, with increasing levels accompanying the increase in the severity of the disease. Gal-10 is a hallmark of eosinophil death and suffers a phase transition to a crystalline state, originating the CLCs that promote type 2 immunity. The induction of type 2 immunity is related specifically to the crystalline state of Gal-10 and is not seen with its soluble form. Moreover, CLCs have proinflammatory effects in the lungs, leading to an influx of neutrophils and monocytes after 6 and 24 h, respectively, after injection, which is accompanied by the production of large quantities of IL-6 and TNF-α in bronchoalveolar lavage fluid and of IL-1β and of CCL2 (monocyte chemoattractant) in lung tissue. Furthermore, CLCs boost adaptive immunity to co-administered antigens (57). Thus, the increase in Gal-10 in critical and severe patients, in a dose-response fashion, might be associated with the “cytokine storm” that is characteristic of the poorer outcomes of COVID-19. Recently, a pre-clinical study showed that antibodies against Gal-10 to preformed CLCs completely dissolved the crystals present in the mucus within 2 hours (57). These antibodies should be evaluated as a potential therapy for SARS-CoV-2 positive patients that are diagnosed with high levels of CLCs at admission (**Table 6**).

According to our findings, potential therapies to COVID-19 might include supplementation with plasma form GELS, increasing the expression of POLR3D in initial stages of infection, administration of antibodies against CLCs, increasing the expression of IREB2, increasing pulmonary surfactant-associated proteins, increasing in PON1 and modulation of the interaction between ULBP6/NKG2D receptor (**Table 6**).

In conclusion, our results indicate several plasma proteins involved in the pathogenesis of COVID-19 that might be useful to predict the prognosis of the disease, when analyzed upon admission of the patients to hospital. These proteins are mainly associated with inflammation, immune response, and blood coagulation. Targeting their pathways might constitute potential new therapies for the disease, which should be evaluated in further studies. Validation of some of the identified candidates is currently being conducted. Analyses of plasma samples collected from these patients at multiple time points until discharge or death are underway, which will allow more controlled longitudinal analysis of severity.

## Data Availability

Upon request, the corresponding author may supplu the data. Additional analyses are still being conducted. After they are finished, all data will be deposited at the repository of the University of Sao Paulo.

http://dadoscientificos.usp.br

## Acknowledgements

We would like to thank State of São Paulo Research Foundation (FAPESP) for the financial support through the Thematic (#2015/03965-2) and Regular (#2018/12041-7) Projects, the National Council for Scientific and Technological Development (CNPq, #302371/2018-4 and #307986/2017-9) and Coordenação de Aperfeiçoamento de Pessoal de Nível Superior – Brasil (CAPES) – Finance Code 001. The funders had no role in study design, data collection and analysis, decision to publish or preparation of the manuscript. All authors gave their final approval and agree to be accountable for all aspects of the work. The authors thank Prof. Daniel Martins de Souza for providing advice on plasma proteomic analysis.

